# Economic analysis of the impact of overseas and domestic treatment and screening alternatives for parasitic infections among U.S.-bound refugees from Sub-Saharan Africa, 2013-2022

**DOI:** 10.64898/2026.09.04.26358165

**Authors:** Brian Maskery, Heesoo Joo, Joohyun Park, Michelle Weinberg, Tarissa Mitchell, Alexander Klosovsky, William M. Stauffer

## Abstract

U.S.-bound refugees from sub-Saharan Africa arrive from countries where parasitic infections are highly endemic (including hookworm, ascariasis, trichuriasis, strongyloidiasis, schistosomiasis, and malaria). These infections are rare in the United States and may be underdiagnosed or misdiagnosed. This evaluation examined costs and health outcomes of four alternatives: 1) “Overseas Presumptive Treatment”, 2) “Domestic Screening and Treatment”, 3) “Domestic Presumptive Treatment”, and 4) “No Program” using decision trees terminating in Markov transition state models. The current program, “Overseas Presumptive Treatment”, is the least expensive and most effective alternative with baseline parameter estimates. Relative to “No Program”, the net cost per QALY gained for “Overseas Presumptive Treatment” has 95% uncertainty interval (UI) from cost-saving to $52,000, Domestic Screening and Treatment”: 95% UI: $6,700-$580,000, and “Domestic Presumptive Treatment”: 95% UI $12,000-$1.29 million. Even using conservative parameter assumptions, overseas presumptive treatment is cost-effective and reduces the burden of parasitic infections among sub-Saharan refugees.

**Article Summary Line:** In this analysis, we estimated the costs and health outcomes of CDC’s presumptive treatment program for parasitic diseases for U.S.-bound refugees from sub-Saharan Africa and compared these estimates to three hypothetical alternatives.

## Background

The total number of refugees resettling in the United States varied between 2013 and 2022 from a low of 11,454 in 2021 and a high of 84,989 in 2016.(1) The annual number of refugees resettled from Africa averaged 15,656 and varied from 4,173 to 31,640 over the ten-year period.(1) Refugees resettling in the United States have been reported to have high prevalence rates of parasitic infections.(2–6) These diseases are rare in the United States, which may lead to errors or delays in diagnosis or inappropriate treatment.(7–9)

In 1997, the U.S. Centers for Disease Control and Prevention (CDC) initiated overseas presumptive treatment for refugees enrolled in the U.S. Refugee Admissions Program (USRAP).(10) The program initially included a single dose of albendazole for intestinal nematodes to all refugees departing from Asia and Africa and sulfadoxine-pyrimethamine for anti-malarial treatment among refugees departing from sub-Saharan Africa. For U.S.-bound refugees from (sub-Saharan) Africa, the program has evolved to include four drugs (albendazole, ivermectin, praziquantel, and artemether-lumefantrine) targeting six parasitic infections (hookworm, trichuriasis, ascariasis, strongyloidiasis, schistosomiasis, and *P. falciparum* malaria). (11) The full CDC guidance is available online.(10) Most infections may be asymptomatic or associated with vague clinical symptoms including abdominal complaints, nausea, diarrhea, constipation, or skin manifestations.(12) Strongyloidiasis and malaria can cause severe disease requiring hospitalization or be fatal.(8, 9, 13) Presumptive treatment is administered shortly before departure to the United States to minimize the risk of re-infection.

Previous work suggests presumptive treatment of parasitic infections among U.S.-bound refugees and immigrants from high-burden countries is cost-effective.(4, 14–16). However, previous analyses had limited or outdated data on U.S. costs of treating these diseases. Recent studies provided updated U.S. treatment cost estimates and have found low utilization of appropriate anti-parasitic drugs for individuals treated as outpatients.(17–21) Our objective was to quantify and compare the benefits and costs of overseas presumptive parasite treatment with domestic screening and treatment, domestic presumptive treatment and no program for the refugee population resettling from sub-Saharan Africa.

## Methods

We developed a decision tree model to assess the costs and health impacts of four alternatives: “No Program“; “Overseas Presumptive Treatment”; “Domestic Screening and Treatment”; and “Domestic Presumptive Treatment”.

For each alternative, refugees were subdivided according to whether they were infected with one of the six parasites or not infected. We did not account for simultaneous infections due to lack of data on the severity of multiple versus single infections. The fraction of refugees infected with each parasite prior to intervention is based on published prevalence estimates.(4, 6, 14, 22–29) More details on parameter assumptions are included in Tables S1 through S22 of Supplemental Information. In general, uncertain parameters, such as QALY decrements, were estimated to result in conservative health outcomes from untreated infections.

Under the “No Program” alternative, refugees do not undergo any intervention and only receive treatment if they present with illness. The only costs are associated with diagnosis and treatment if refugees present for treatment after arrival. As summarized in S1 and S22, the probability of seeking treatment if infected is relatively low across all six infections.

“Overseas Presumptive Treatment” is the current program (11) where refugees are presumptively treated with albendazole, ivermectin, praziquantel, and artemether-lumefantrine overseas before departing for the United States. Under this alternative, some fraction (10%) of refugees may not receive presumptive treatment due to ineligibility (e.g., contraindications including pregnancy). Included in this 10% were refugees who originated from or had lived in *Loa loa*-endemic areas who do not receive presumptive treatment for strongyloidiasis (due to risk of encephalitis if a patient has a high *Loa loa* microfilarial load).(30) Ineligible individuals have a second chance to undergo domestic screening and treatment (as described below) after arrival. The model includes potential treatment failures based on drug-specific effectiveness. Drug efficacies were assumed to be similar overseas and in the United States; a multiplier of 0.95 (range: 0.90 to 1.0) was used to adjust overseas treatment efficacy.

Under “Domestic Screening and Treatment”, presumptive treatment would not be provided prior to departure. Instead, refugees would receive a battery of recommended tests after arrival at their domestic comprehensive medical examinations and receive treatment if indicated.(10) We assumed that 10% of refugees would not present for comprehensive examinations and this group would have the same likelihood of developing symptomatic illness as those in the “No Program” alternative. Our analysis also includes consideration of both false positive (only for strongyloidiasis, malaria, and schistosomiasis) and false negative (for all parasites) screening test results based on reported specificities and sensitivities of each test (Tables S1 and S22). If refugees received false positive results, we assumed they would incur costs associated with follow-up and drug treatment despite not being infected. If they received false negative results, they would not be treated and would remain infected.

Finally, “Domestic Presumptive Treatment” modeled presumptive treatment of eligible refugees after U.S. arrival with the same four-drug drug regimen as “Overseas Presumptive Treatment”. As above, we assumed that a fraction of refugees would not present for the domestic comprehensive examination and would not be treated and that another fraction of refugees who do present for the examination instead undergo domestic screening and treatment due to contraindications or other concerns.

The decision tree quantified fractions of infected vs. uninfected individuals by disease. Then, we used Markov transition state models to estimate transition to outpatient treatment, inpatient treatment, uninfected status, or death and to estimate the number of years spent with parasitic infections. We assumed that refugees died of causes unrelated to parasitic infections at rates equivalent to the age-specific U.S. population. Infected individuals transitioned to uninfected status without treatment based on the average organism lifespan (duration of human infection) by parasite or if they were treated as inpatients or outpatients. Hospitalized patients could die based on estimated case fatality rates for malaria and strongyloidiasis., We applied quality-adjusted life year (QALY) weights from the U.S. population by age.(31) Except for individuals with schistosomiasis infections, we subtracted a small QALY decrement (0.001) for each year spent with infection, consistent with a previous analysis.(4) Individuals who required hospitalization for either strongyloidiasis (0.0054) or malaria (0.0036) were assumed to have higher QALY decrements for the period in which they would be hospitalized (Table S21). We applied QALY decrement of 0.006 for individuals with schistosomiasis infections.(4, 32, 33) This decrement was used in lieu of attempting to estimate disutility associated with potential sequela associated with chronic schistosomiasis including renal failure, stroke, or infertility, which have been investigated by other authors.(33–35) Sensitivity analyses evaluated uncertainty in these decrements.

The average age and annual number of African refugee arrivals were based on data from the Department of Homeland Security from 2013 through 2022.(1) Domestic screening and treatment and presumptive treatment costs were estimated as described in Supplemental Information (Tables S9 – S16, S19, and S22).(17–19, 21) Overseas presumptive treatment costs were estimated using the fiscal year 2023 budget of the International Organization for Migration (IOM) which was the primary overseas contracted health service provider for U.S.-bound refugees. We discounted costs and health outcomes by 3% annually over a period of 65 years after arrival.

For each alternative, we calculated total program costs (incurred in the first year). We also calculated the total number outpatient cases, hospitalizations, and deaths associated with parasitic infections and the number of QALYs for the population. Discounted healthcare costs for outpatient and inpatient treatment were added to program costs. Incremental cost-effectiveness ratios (ICERs) were presented as net costs per case, hospitalization, and death averted or QALY gained. ICERs were calculated from sums of program costs (***C****_program_i_*) and discounted illness costs (***C****_illness_i_*) for each alternative (*i*): [***C****_program*_2_ + ***C****_illness*_2_] - [***C****_program*_1_ + ***C****_illness*_1_] divided by differences in cases, hospitalizations, deaths, or QALYs (e.g., [*cases*_2_ + *cases*_1_] - [*QALYs*_2_ + *QALYs*_1_] where subscripts refer to one of the four alternatives. Net cost calculations do not account for lost productivity or reduced consumption due to premature death since refugees may differ from the general population.

To account for uncertainty, we conducted one-way, two-way, and probabilistic sensitivity analyses of ICER estimates. One-way sensitivity analyses were conducted by varying individual parameters across their uncertainty ranges while holding all other parameters at baseline values. Two-way sensitivity analysis is shown in Supplemental Information. We used Monte Carlo Simulation to randomly draw 50,000 samples of uncertain parameters using probability distributions summarized in Tables S1 and S9 for multivariate probabilistic sensitivity analysis. We used Microsoft Excel (Redmond, WA) with the Lumivero @Risk Excel plugin (Denver, CO). This activity used previously collected aggregate data and did not involve contact with human subjects. This activity was reviewed by CDC, deemed research not involving human subjects, and was conducted consistent with applicable federal law and CDC policy.^§^

## Results

### Health impact and costs

The “No Program” alternative would cost $980,073 per cohort of 15,656 U.S.-bound refugees from Sub-Saharan Africa based on the annual average number of refugee arrivals from 2013-2022 (Table 1). Program costs are zero and only outpatient (all six parasites) or inpatient strongyloidiasis and malaria cases incur costs. The “Overseas Presumptive Treatment” alternative ($873,063), which is the current program, is less costly than “Domestic Screening and Treatment” ($3,590,905) or “Domestic Presumptive Treatment” ($6,727,753). “Overseas Presumptive Treatment” includes overseas presumptive treatment program costs ($412,841) for 90% of individuals, costs for domestic follow-up of the 9% of individuals who would be unable to receive presumptive treatment ($341,297) and illness costs incurred after completing the intervention ($118,926). For “Domestic Screening and Treatment”, screening costs ($1,578,095) are less than treatment costs for those diagnosed with infections ($1,834,874).

**Table 1.** Estimated baseline costs and health outcomes for parasite control alternatives among U.S.- bound Sub-Saharan Africa n refugees, 15,656 annual cohort.

|  | “No Program” | “Domestic Screening and Treatment” | “Overseas Presumptive Treatment” (Current program) | “Domestic Presumptive Treatment” |
| --- | --- | --- | --- | --- |
| Fraction of target population undergoing domestic screen/treat | 0% | 90% | 9% | 9% |
| Fraction of target population undergoing overseas (pre-departure) presumptive treatment | 0% | 0% | 90% | 0% |
| Fraction of target population undergoing post-arrival (domestic) presumptive treatment | 0% | 0% | 0% | 81% |
| Fraction of target population with no intervention | 100% | 10% | 1% | 10% |
| **Program (intervention) costs** |  |  |  |  |
| Overseas or domestic presumptive treatment costs | \$0 | \$0 | \$412,841 | \$6,226,428 |
| Domestic screening costs | \$0 | \$1,437,193 | \$143,719 | \$143,719 |
| Domestic treatment costs to treat individuals identified during domestic screening | \$0 | \$1,768,083 | \$176,808 | \$176,808 |
| Opportunity costs for domestic screening | \$0 | \$140,901 | \$14,090 | \$14,090 |
| Opportunity costs to treat individuals diagnosed during domestic screening | \$0 | \$66,791 | \$6,679 | \$6,679 |
| Total costs to complete intervention | \$0 | \$3,412,968 | \$754,138 | \$6,567,725 |
| Average intervention cost per refugee <sup>a</sup> | \$0 | \$218 | \$48 | \$420 |
| **Markov model: illness cost estimates <sup>b</sup>** |  |  |  |  |
| Estimated discounted treatment costs <sup>b</sup> | \$914,971 | \$166,581 | \$111,351 | \$149,708 |
| Estimated discounted illness opportunity costs <sup>a</sup> | \$65,103 | \$11,356 | \$7,574 | \$10,319 |
| Total discounted illness costs <sup>b</sup> | \$980,073 | \$177,937 | \$118,926 | \$160,027 |
| **Total costs (intervention + illness costs) <sup>b</sup>** |  |  |  |  |
| Total intervention + illness costs | \$980,073 | \$3,590,905 | \$873,063 | \$6,727,753 |
| (including illness opportunity costs)<sup>b</sup> |  |  |  |  |
| Total intervention + illness costs (excluding illness opportunity costs)<sup>b</sup> | \$914,971 | \$3,579,549 | \$865,489 | \$6,717,433 |
| Number of individuals infected (after intervention) |  |  |  |  |
| Hookworm | 282 | 127 | 77 | 87 |
| Ascariasis | 1,660 | 492 | 174 | 246 |
| Trichuriasis | 1,205 | 871 | 841 | 859 |
| Strongyloidiasis | 1,706 | 520 | 362 | 427 |
| Schistosomiasis | 1,425 | 689 | 498 | 543 |
| Malaria | 1,127 | 148 | 94 | 144 |
| Total | 7,405 | 2,846 | 2,047 | 2,305 |
| Markov model: health outcomes |  |  |  |  |
| Total outpatient cases | 114 | 34 | 24 | 28 |
| Total hospitalized cases | 34.5 | 5.0 | 3.2 | 4.7 |
| Total deaths | 0.64 | 0.16 | 0.11 | 0.13 |
| Total discounted QALYs<sup>b</sup> | 314,579 | 314,639 | 314,649 | 314,646 |
| Net cost estimates and incremental cost-effectiveness ratio estimates |  |  |  |  |
| Net costs relative to "No program" (including illness opportunity costs) | NA | \$2,610,832 | (\$107,010) | \$5,747,679 |
| Outpatient cases averted | NA | 79.8 | 90.0 | 85.7 |
| Hospitalizations averted | NA | 29.5 | 31.3 | 29.8 |
| Deaths averted | NA | 0.49 | 0.53 | 0.51 |
| QALY gained | NA | 60 | 70 | 67 |
| Net cost per case averted | NA | \$17,520 | (\$747) | \$38,624 |
| Net cost per hospitalization averted | NA | \$88,434 | (\$3,422) | \$192,988 |
| Net cost per death averted | NA | \$5,372,114 | (\$200,072) | \$11,286,203 |
| Net cost per QALY gained | NA | \$43,535 | (\$1,520) | \$85,774 |

<sup>a</sup> Intervention cost per refugee is calculated from the intervention cost divided by the cohort size (n= 15,656).
<sup>b</sup> Illness cost estimates are discounted at 3% per year over a period of 65 years. QALYs are also discounted at 3% over the same period. Cases, deaths and hospitalizations were not discounted.

After completing each alternative, the numbers of refugees expected to remain infected with any parasite would be 7,405 for “No Program”, 2,846 for “Domestic Screening and Treatment”, 2,047 for “Overseas Presumptive Treatment”, and 2,305 for “Domestic Presumptive Treatment” (Table 1). A relatively small fraction of infected individuals would present with clinical disease: up to 114 outpatient cases, 34.5 hospitalizations, and 0.64 deaths attributable to these parasitic infections over 65 years for “No Program”. The remaining infected individuals were assumed to clear the infection without treatment based on the expected duration of infection or to die of other causes. “Overseas Presumptive Treatment” would avert 90 outpatient cases, 31 hospitalizations, and 0.51 deaths, and result in 70 QALYs gained relative to “No Program”. “Domestic Screening and Treatment” would avert 80 outpatient cases, 30 hospitalizations, and 0.49 deaths, and result in 60 QALYs gained. “Domestic Presumptive Treatment” would avert 86 outpatient cases, 30 hospitalizations, and 0.51 deaths, and result in 67 QALYs gained. Patients with strongyloidiasis comprised about 75% of outpatient cases and 64% to 82% of deaths depending on alternative. Patients with malaria comprised between 85% and 93% of those hospitalized (Table S23 in Supplemental Information).

For the baseline analysis, “Overseas Presumptive Treatment” is the dominant alternative because it is both less expensive and results in better health outcomes relative to “No Program”, “Domestic Screening and Treatment”, or “Domestic Presumptive Treatment.” The net cost per QALY gained are estimated at $43,535 for “Domestic Screening and Treatment” and $85,774 for “Domestic Presumptive Treatment” relative to “No Program”.

### Sensitivity analysis

Tornado diagrams demonstrate how varying one parameter at a time for the 20 most influential parameters impact ICER estimates (net cost per QALY gained) for three alternatives relative to “No Program”. “Overseas Presumptive Treatment” was found to be cost-saving with base case parameter estimates (Table 2); however, there were ten individual parameter estimates for which it would not be considered cost-saving. ICER estimates were less than $5,000 across all parameters. The malaria prevalence, cost of inpatient malaria treatment, and annual probability of inpatient treatment for malaria were the most influential individual parameters (Figure 1a). ICER estimates for “Domestic Screening and Treatment” relative to “No Program” varied across a range of between $8,000 and $91,000 according to variation in individual parameter estimates (Figure 1b). The most important parameter estimates influencing ICER estimates are 1) subclinical or outpatient strongyloidiasis QALY decrement, 2) strongyloidiasis prevalence, and 3) schistosomiasis prevalence. The ICER estimates for “Domestic Presumptive Treatment” relative to “No Program” are highest and vary from $16,000 to $200,000 across one-way sensitivity analyses (Figure 1c). Figures S4a-c in Supplemental Information present tornado diagrams including uncertainty across all parameter estimates. Two-way sensitivity analyses are presented in Tables S22a-c in Supplemental Information.

**Figure 1a.**
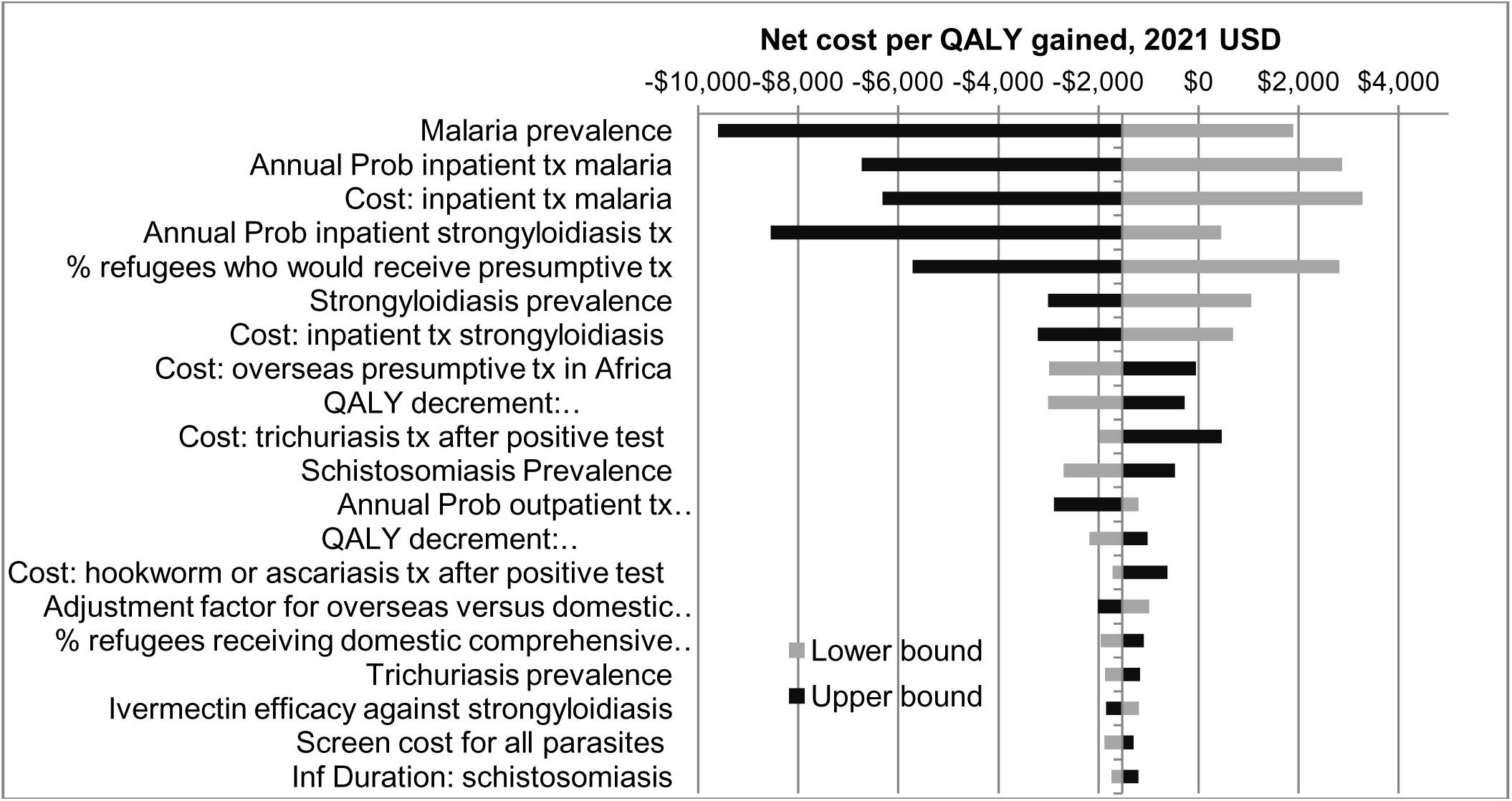
One-way sensitivity analysis of ICER estimates (net cost per QALY gained) for “Overseas Presumptive Treatment” (current program) vs. “No Program”, top 20 parameters causing the greatest difference in ICER estimates between lower and upper bounds, base case value: -$703 2021 USD

**Figure 1b.**
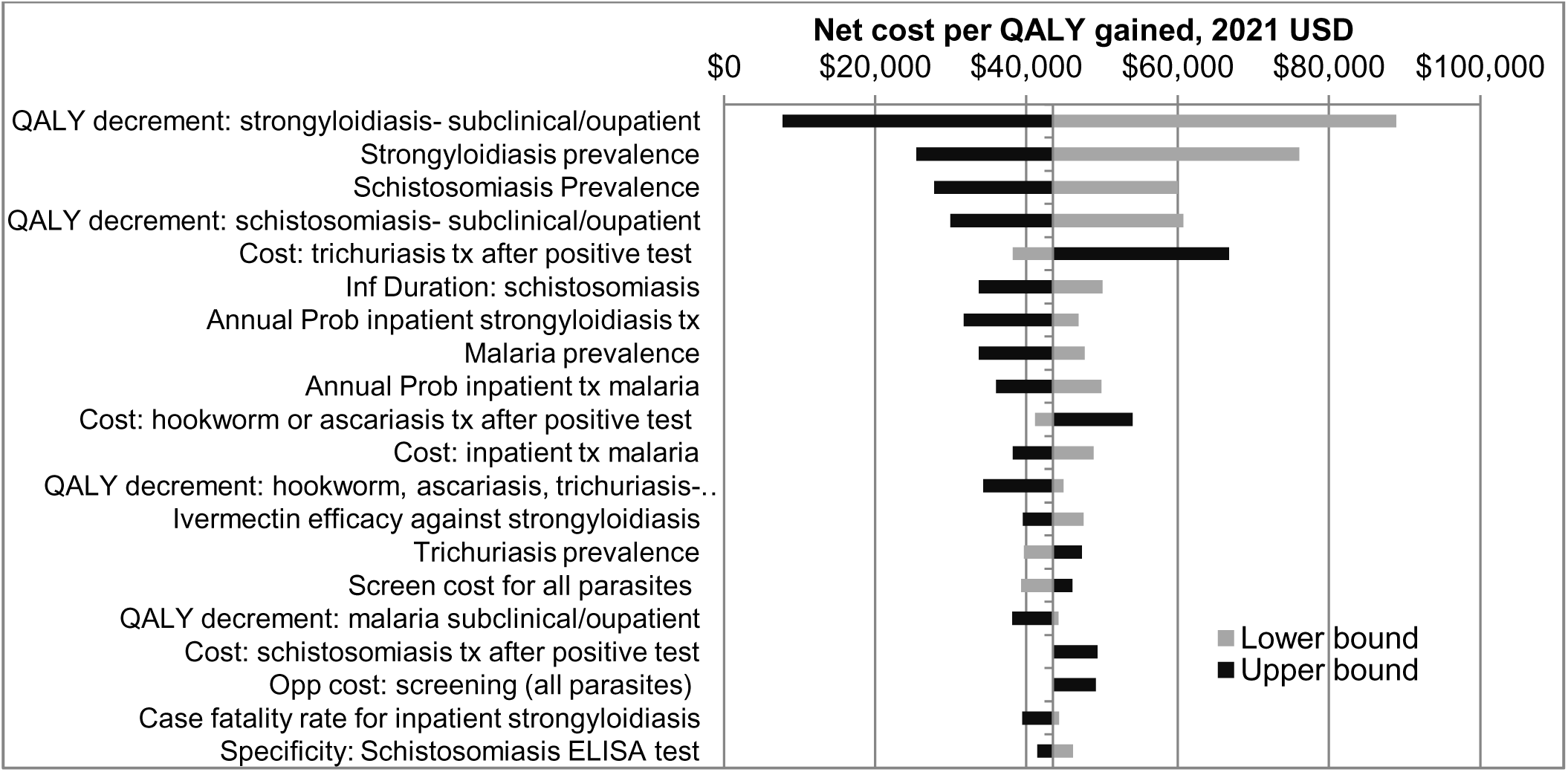
One-way sensitivity analysis of ICER estimates (net cost per QALY gained) for “Domestic Screening and Treatment” vs. “No Program”, top 20 parameters causing the greatest difference in ICER estimates between lower and upper bounds, base case value: $44,431, 2021 USD

**Figure 1c.**
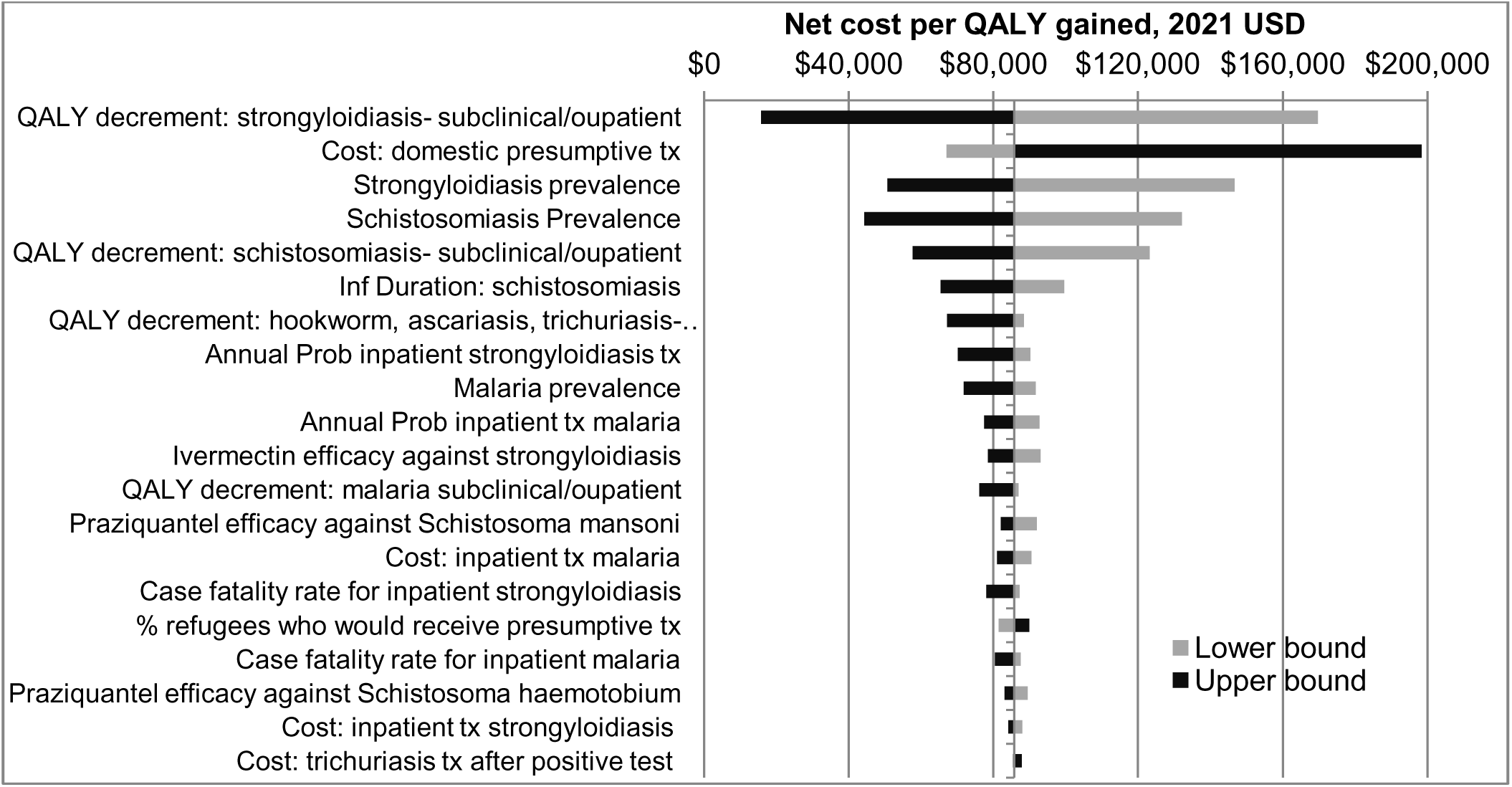
One-way sensitivity analysis of ICER estimates (net cost per QALY gained) for “Domestic Presumptive Treatment” vs. “No Program”, top 20 parameters causing the greatest difference in ICER estimates between lower and upper bounds, base case value: $86,591, 2021 USD

Uncertainty intervals (95%) were estimated from the results of the multivariate probabilistic sensitivity analysis using Monte Carlo Simulations. Relative to “No Program”, the net cost per QALY gained ICER varies from cost saving to $52,000 for “Overseas Presumptive Treatment” (baseline value: cost-saving), from $6,900 to $590,000 for “Domestic Screening and Treatment” (baseline value: $44,431), and from $12,000 to $1.3 million for “Domestic Presumptive Treatment” (baseline value: $86,591).

Figure 2 presents the fraction of Monte Carlo Simulation iterations below a given willingness to pay threshold for gaining one QALY. “Overseas Presumptive Treatment” was found to result in cost-savings in about 45% of the iterations, and ICER estimates of less than $10,000, $50,000, $100,000 or $200,000 in 80%, 97%, 99.5%, or 99.9% of the iterations, respectively, relative to “No Program”. In comparison, “Domestic Screening and Treatment” had ICER estimates of less than $10,000, $50,000, $100,000 or $200,000 in only 5%, 28%, 52%, or 77% of iterations, respectively. Finally, “Domestic Presumptive Treatment” is the least likely to be cost-effective for any given willingness to pay threshold. Figure S5 in Supplemental Information provides a log-scale figure. Figures S6a-8b provide scatter plots of the net cost and net QALYs gained for each intervention alternative relative to “No Program”.

**Figure 2.**
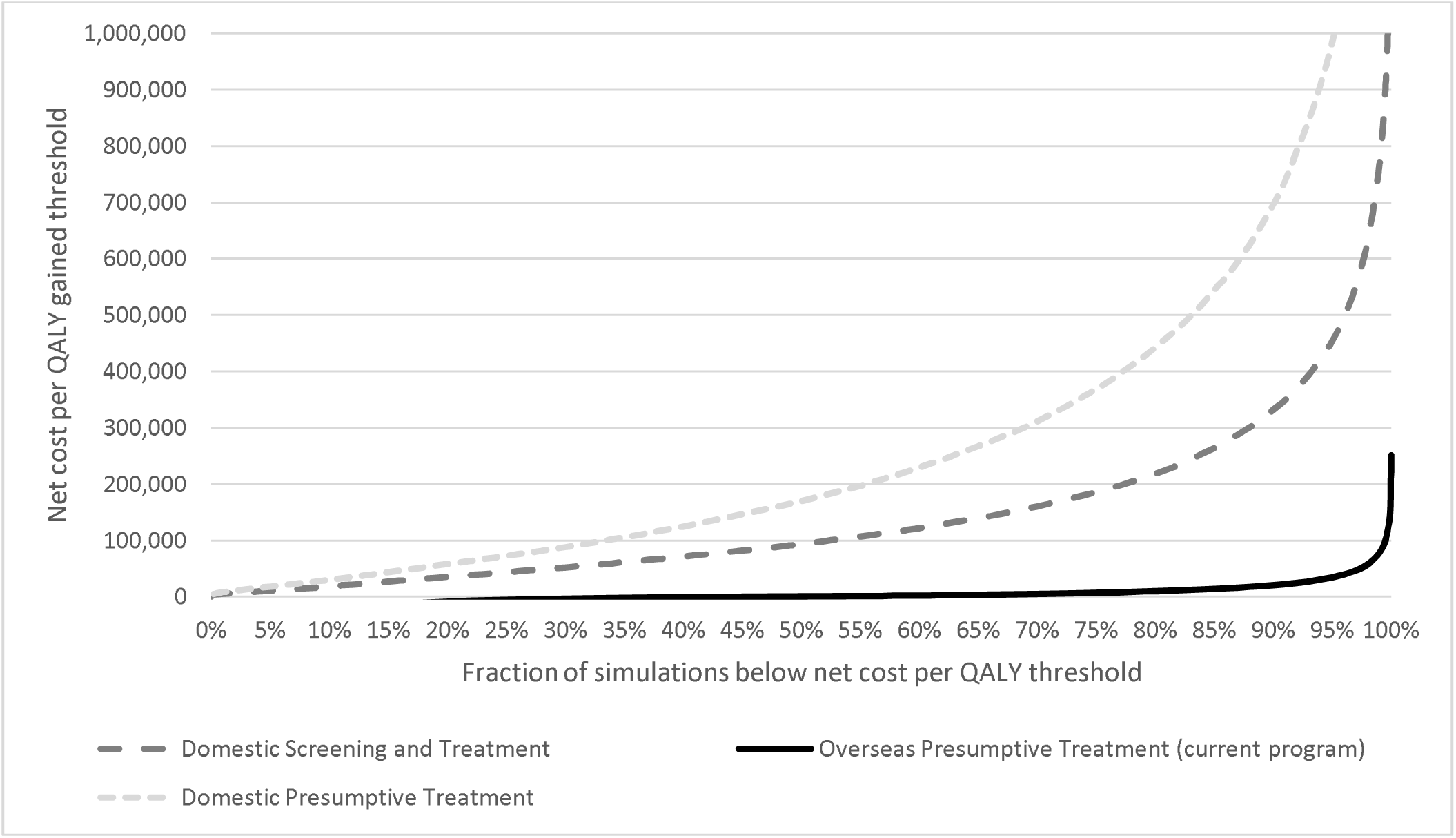
Fraction of Monte Carlo Simulation iterations in which each intervention would be considered cost-effective as a function of willingness to pay per QALY gained for “Overseas Presumptive Treatment”, “Domestic Screening and Treatment”, or “Domestic Presumptive Treatment” relative to “No Program”

## Discussion

CDC’s current program (11) is to provide overseas presumptive treatment with four drugs (albendazole, ivermectin, praziquantel, and artemether-lumefantrine) to reduce the burden of six parasites (hookworm, ascariasis, trichuriasis, strongyloidiasis, schistosomiasis, and *P. falciparum* malaria). Our analysis indicates “Overseas Presumptive Treatment” is a cost-effective intervention for improving the health of U.S.-bound refugees from Sub-Saharan Africa prior to arrival in the United States. Although there is considerable uncertainty in our parameter estimates, the net cost per QALY gained ICER remained below $10,000 in 80% of our simulations compared to waiting for symptomatic U.S.-bound refugees to present with illness in the United States (i.e., “No Program”). Compared with “Domestic Screening and Treatment” or “Domestic Presumptive Treatment”, “Overseas Presumptive Treatment” is both less expensive and more effective. Overseas presumptive treatment can be accomplished for large groups of U.S.-bound refugees, a less complex process than post-arrival follow-up. In general, overseas presumptive treatment includes lower labor and drug costs, can result in higher treatment completion rates (Appendix Tables S7 and S9), and less U.S. provider education. (4)

This finding, that overseas presumptive treatment of U.S.-bound refugees from Sub-Saharan Africa is cost-effective, aligns with findings from an analysis conducted for U.S.-bound refugees from Asia who undergo presumptive treatment with albendazole and ivermectin (4), as well as findings from an analysis involving West African refugees presumptively treated for malaria. (14) Our findings are also consistent with findings from cost-effectiveness studies (15, 16) of the presumptive use of albendazole and ivermectin for individuals in the United States who immigrated from countries where hookworm, ascariasis, trichuriasis, and strongyloidiasis are endemic. This analysis builds upon previous analyses by including presumptive treatment for schistosomiasis, which had not been studied previously among U.S.-bound refugees. Our results are also consistent with European studies that showed presumptive treatment for strongyloidiasis (after arrival in Europe for these studies) would be cost-effective and may be cost-saving if taking account of potential misdiagnoses leading to higher treatment costs over time.(36, 37) The analysis is unique in its consideration of four drugs for six parasitic diseases. By combining interventions for multiple diseases, the program should be more efficient relative to interventions targeting a single parasitic disease since the drugs are administered at the same time.

This analysis includes updated treatment costs for these six diseases, which incorporate the increased U.S. prices of antiparasitic drugs (especially albendazole and praziquantel) in recent years. (17–21) These increasing drug prices reduce the cost effectiveness of treating individuals presumptively after arrival or treating individuals who would test positive during a domestic screening program. We assumed that 90% of individuals under “Overseas Presumptive Treatment” would be treated prior to departure at an average cost per refugee to receive of $29.30. We further assumed that 9% would be unable to complete presumptive treatment overseas and incur costs for domestic screening and treatment instead (average $218 per person), resulting in a higher overall cost of $48 per refugee than if all refugees had been presumptively treated. “Overseas Presumptive Treatment” becomes more cost effective as the percentage of individuals treated overseas increases.

This analysis has limitations. First, since the United States has been providing overseas presumptive treatment to refugees for many years, our prevalence estimates rely on older data (6) or on data that may not be specific to or representative of U.S.-bound refugee populations. For instance, the prevalence of schistosomiasis (9.1%) and strongyloidiasis (10.9%) were estimated from a systematic review of rates in migrants. (22) The prevalence of malaria (7.2%) was estimated from studies conducted in many countries. (14, 23–29) [Table S4] The actual prevalence of each infection is likely to vary each year depending on the distribution of countries from which refugees arrive.

Next, the QALY burden associated with chronic parasitism is difficult to quantify, especially in people outside an endemic area. Most people would not be aware that they were infected and would have non-specific symptoms from conditions associated with infection (e.g., anemia with hookworm, abdominal complaints with strongyloidiasis or schistosomiasis). Prior to implementing presumptive treatment for U.S.-bound refugees from Sub-Saharan Africa, high prevalence rates of schistosomiasis and strongyloidiasis were reported in refugees from Sudan and Somalia who underwent post-arrival testing; however, individuals who tested positive did not report statistically different symptoms than individuals who tested negative. (5) We used conservative QALY decrements in this analysis to account for the potetital subclinical nature of these infections. In addition, we conducted additional sensitivity analyses across multiple parameters (Table S22a-c, Figures 1a, 1b, and 2) that showed the cost per QALY gained would remain below $10,000 across a range of parameter estimtates.

Third, although this study focuses on six parasitic infections, presumptive treatment with albendazole, ivermectin, and praziquantel would impact other infections such as enterobiasis, clonorchiasis, and tapeworms among others. Previous studies have indicated low usage of standard-of-care treatment for these infections in the United States. (38) Thus, the benefit of presumptive treatment is likely underestimated.

We assumed that individuals treated as outpatients or inpatients would clear their infections in the Markov model. This approach differed from our assumption in the decision tree model that individuals that undergo presumptive treatment or domestic screening may have drug treatment failures. We believe this assumption in the Markov model is partly justified by the time-limited duration of most of these infections (i.e., even if drug treatment does not eliminate the parasitic infection in some individuals, the reduction in the intensity of infection may be sufficient to minimize the infection-related disutility for the remainder of each parasite’s life cycle). However, strongyloidiasis may persist indefinitely in individuals for whom drug treatment is ineffective. This approach would underestimate the disutility caused by strongyloidiasis across all interventions including “No Program”.

Due to complexity and to be conservative, we did not include the cost and health impact of long-term schistosomiasis-associated sequela including ascites, bladder cancer and others. (35) Our use of conservative QALY decrement estimates and lack of consideration for schistosomiasis-associated long-term sequela would underestimate the number of QALYs gained and overestimate ICERs for any of the intervention alternatives relative to “No Program”. We also did not attempt to adjust for the cost of treating malaria using intravenous artesunate, which was recently approved for use in the United States, replacing the use of either the lower cost intravenous quinidine or intravenous artesunate provided through CDC under an investigational new drug protocol.(39) Commercial availability of intravenous artesunate has increased the cost of treating severe malaria but this analysis used treatment cost data preceding commercial availability. As discussed further in Supplemental Information, we made several conservative assumptions regarding the likelihood of developing symptomatic malaria among infected individuals that could underestimate the cost of ‘No Program’ relative to other alternatives considered in this analysis.

We estimated the costs, screening test sensitivities, and test specificities by assuming that refugees would be screened in accordance with the CDC refugee parasite guidance. Domestic examinations may include fewer, or more likely, more extensive diagnostic testing. Finally, we had a limited number of studies available to extrapolate outpatient and inpatient cases resulting from infections. (4, 15, 16, 40) More complete surveillance data are only available for U.S. malaria patients.(13, 41).

## Conclusions

The high cost of drugs, screening and diagnostic tests, and logistics of follow-up care and limited provider knowledge about neglected parasitic diseases in the United States create an environment where overseas presumptive parasite treatment for U.S.-bound refugees from Sub-Saharan Africa is the most cost-effective approach to reducing the burden of parasitic infections among refugees resettling to the United States. Overseas presumptive treatment remains cost-effective even using conservative assumptions of health outcomes associated with untreated infections and improves the health of refugees by reducing their parasite burden and hospitalizations due to malaria and strongyloidiasis. Presumptive treatment also protects the public health of the United States by minimizing any potential exposures to these infections.

## Data Availability

All data are available upon reasonable request to the authors.

## Acknowledgments

The findings and conclusions in this report are those of the authors and do not necessarily represent the official position of the Centers for Disease Control and Prevention or the institutions with which the authors are affiliated.

## Supplemental Information

S0. Online Appendix

## Footnotes

§ See e.g., 45 C.F.R. part 46; 21 C.F.R. part 56; 42 U.S.C. §241(d), 5 U.S.C. §552a, 44 U.S.C. §3501 et seq.

