## Supplemental appendix for "Economic analysis of the impact of overseas and domestic treatment and screening alternatives for parasitic infections among U.S.-bound refugees from Sub-Saharan Africa, 2013-2022"

^4^ Department of Medicine. Division of Infectious Diseases and International Medicine, University of Minnesota, Minneapolis, MN, USA

*Brian Maskery

Health Economist

Division of Global Migration Health

National Center for Emerging and Zoonotic Infectious Diseases

Centers for Disease Control and Prevention

1600 Clifton Road NE, Mailstop H16-4

Acknowledgments

The findings and conclusions in this report are those of the authors and do not necessarily represent the official position of the Centers for Disease Control and Prevention or the institutions with which the authors are affiliated.

### Decision Tree and Markov models

*Decision tree models*

The decision tree models used in this analysis were developed similar to the approach used in Maskery et al. (2016) (1). The costs and health impacts of alternative programs were estimated using a decision tree model terminating with Markov processes. Each individual considered in the model could either be infected with exactly one parasite (causing ascariasis, trichuriasis, hookworm, strongyloidiasis, schistosomiasis, or malaria) or be uninfected at the initiation of the Markov process. A simplified schematic of the decision tree model is shown in Figure S1. The complete decision tree model is available from the corresponding author. For each of the four alternatives under consideration (i.e., “No Program”, “Domestic Screening and Treatment”, “Overseas Presumptive Treatment”, and “Domestic Presumptive Treatment”), refugees begin in either an infected (with one of the six parasites) or uninfected state. For “No Program”, individuals would exit the decision tree and enter the Markov model in either the disease-specific infected state or uninfected state based on the estimated prevalence rates. For the three intervention alternatives: “Domestic Screening and Treatment”, “Overseas Presumptive Treatment”, or “Domestic Presumptive Treatment”, individuals may participate in the intervention opportunity. In addition, for “Overseas Presumptive Treatment”, individuals may not have access to the program in the country from which they depart because of certain health conditions (e.g., pregnancy) or may choose not to participate. Those individuals would be eligible to instead undergo “Domestic Screening and Treatment” after arrival in the United States. We assumed that refugees who receive presumptive treatment prior to departure would not be screened for parasitic diseases after arrival in the United States. If individuals do not receive overseas presumptive treatment and do not follow up for domestic screening and treatment, they would all be considered untreated, and their risk of disease progression would be the same as for individuals under “No Program”. The “Domestic Presumptive Treatment” alternative is like “Overseas Presumptive Treatment” except that presumptive treatment occurs after arrival rather than prior to departure. In addition, we made parallel assumptions regarding individuals ineligible for treatment or who may choose not to participate. Unlike “Overseas Presumptive Treatment”

When considering “Domestic Screening and Treatment,” some individuals may receive treatment even though they are not infected depending on test specificity. Other individuals may not receive treatment even though they are infected depending on test sensitivity. Both test sensitivity and specificity vary by parasite. The four potential test outcomes are:

1. True Positive: Refugees are infected and treated.
2. True Negative: Refuges are not infected and not treated.
3. False Positive: Refugees are not infected but are still treated.
4. False Negative: Refugees are infected but not treated.

When patients are true positive or false positive, they are assumed to be treated and incur the cost of prescription medications. If they are infected (true positive), the infection will be eliminated at the pathogen-specific efficacy listed in Table 1. If they are not infected (false positive), the refugees would be unnecessarily treated and incur the cost of the medication. If refugees are infected, but test false negative, they will not receive medication and would remain infected when entering the Markov processes described below.

*Markov models*

The Markov model is used to estimate health outcomes for the four alternatives under consideration. For uninfected persons, the Markov process is simple. Each interval, they may die based on background mortality rates in the United States (2). Otherwise, they remain in an uninfected state with quality-adjusted life year (QALY) weight based on the average estimated for the U.S. population (3). The Markov analysis uses a 0.1-year time step and the number of QALYs are evaluated over a 65-year time horizon.

For persons with hookworm, trichuriasis, ascariasis, or schistosomiasis, Figure S2 shows that refugees start in the infected state. Each interval they may: 1) die at the background rate; 2) seek outpatient treatment; 3) remain infected; or, 4) become clear of infection without treatment (based on the assumed duration of infection). We assumed that there are no hospitalizations or deaths caused by these four parasitic diseases. After clearing the infection, we assumed zero probability of reinfection. For each interval spent in the infected state, we assumed a small parasite-specific QALY decrement from the national average QALY weights. We assumed that the infection would clear without treatment after one year on average for ascariasis, two years for trichuriasis, five years for schistosomiasis, and six years for hookworm (Table S1) for the baseline scenario. Both the QALY decrements and duration of infection were varied in sensitivity analyses.

In contrast, individuals with strongyloidiasis or malaria may develop severe disease resulting in hospitalization or death. This adds some additional complexity to the Markov processes considered. A schematic of the Markov model for persons infected with strongyloidiasis or malaria is shown in Figure S3. Each refugee again begins in an infected state. Each iteration, they may 1) remain infected, 2) receive outpatient treatment, 3) receive inpatient treatment, or 4) they may die from other causes at the background mortality rate. If they are treated as inpatients, there is a risk of death (Table 1). We assumed that the infection would clear without treatment after one year on average for malaria, but individuals with strongyloidiasis have lifetime infection without treatment (Table 1) for the baseline alternative. For malaria, our analysis was conducted assuming the vast majority of individuals would be infected with *Plasmodium falciparum*, which constituted 99.3% of identified species among malaria infections in a systematic review of prevalence studies conducted in refugee camps in Africa.{Debash, 2025 #248} Individuals infected with *Plasmodium vivax or Plasmodium ovale* would likely not clear the infection within one year. This is a simplifying assumption to allow for comparison with one year of US surveillance data as described below.

For all six parasitic infections considered in the Markov models, we assumed that any individuals who received outpatient or inpatient treatment would clear the infection unless they died (i.e., we did not consider treatment failures). This is different than the assumptions used for “Domestic Screening and Treatment”, “Domestic Presumptive Treatment” or “Overseas Presumptive Treatment” alternatives evaluated in the initial decision tree model. This is because outpatient cases would occur later after arrival in the United States when infection intensity may be reduced or because more attention may be given to a single infection relative to the comprehensive examination in which multiple health conditions may be addressed.

Figure S1. Simplified schematic of decision tree model

Decision

Overseas Presumptive Treatment

Presumptive treatment program in country and person is elgible

Infected a

Treatment effective (uninfected)

Treatment ineffective (infected) b

Not infected

Refugee treated

No program or not eligibile

Domestic Screening and Treatment.

Domestic Presumptive Treatment

Presumptive treatment program and person is elgible

Infected a

Treatment effective (uninfected)

Treatment ineffective (infected) b

Not infected

Refugee treated

Individual presents for exam but is not eligible

Domestic Presumptive Treatment

Individual does not present for exam

Infected a

Not infected

Domestic Screening and Treatment

Infected a

Follow-up in U.S

True positive

Treatment effective (uninfected)

Treatment ineffective (infected) b

False negative (infected) b

No follow-up (infected) b

Not infected

Follow-up in U.S

False positive

Treatment

True negative

No follow-up

No program

Infected a,b

Not infected

^a^ Persons with one of hookworm, ascariasis, trichuriasis, strongyloidiasis, schistosomiasis, or malaria

^b^ Individuals that complete the decision tree in an infected state, enter the Markov model in an infected state as summarized in Figures S2 and S3. The boxes that are shaded in grey indicate individuals who complete the decision tree process in an infected state.

Figure S2. Schematic of Markov model for hookworm, trichuriasis, schistosomiasis, or ascariasis ^a^

Death (unrelated to parasitic infection)

Infected (asymptomatic or subclinical)

Infection cleared

Outpatient case

Figure S3. Schematic of Markov model for strongyloidiasis or malaria

Death (unrelated to strongyloidiasis or malaria)

Infected (asymptomatic or subclinical)

Infection cleared ^a^

Inpatient case

Outpatient case

Death (strongyloidiasis or malaria)

^a^ Individuals with malaria can clear their infections based on the average duration of infection. Individuals with strongyloidiasis would remain infected unless treated as outpatients or inpatients. The dotted line applies to individuals with malaria, but not individuals with strongyloidiasis.

### The estimation of epidemiological input parameters

The epidemiological input parameters are summarized in Table S1.

Table S1. Epidemiological input parameters

| Description | Baseline | Lower bound | Upper bound | Distri-bution ^a^ | Ref |
| --- | --- | --- | --- | --- | --- |
| **Drug efficacy** | | | | | |
| Albendazole against *Ascariasis* | 0.965 | 0.944 | 0.979 | B | (4) |
| Albendazole against hookworm | 0.785 | 0.715 | 0.842 | B | (4) |
| Albendazole against *Trichuris* | 0.321 | 0.232 | 0.425 | B | (4) |
| Praziquantel against *Schistosoma* mansoni | 0.78 | 0.52 | 0.95 | B | (5) |
| Praziquantel against *S.* haematobium | 0.62 | 0.46 | 0.74 | B | (6) |
| Ivermectin against *Strongyloides stercoralis* | 0.84 | 0.72 | 0.98 | B | (7) |
| Artemether-lumefantrine (AL) against *Plasmodium spp.* (malaria) | 0.97 | 0.96 | 0.98 | B | (8) |
| **Test sensitivity** | | | | | |
| Two stool ova and parasites (O&P) tests for *Ascaris* | 0.81 | 0.57 | 0.95 | B | (1, 9) |
| Two stool O&P tests for hookworm | 0.78 | 0.53 | 0.88 | B | (1, 9) |
| Two stool O&P tests for *Trichuris* | 0.96 | 0.81 | 0.99 | B | (1, 9) |
| *Schistosoma* serologic test (ELISA) | 0.82 | 0.70 | 0.90 | B | (10) |
| *S. stercoralis* serologic test (IVD ELISA) | 0.92 | 0.88 | 0.97 | B | (11) |
| Microscopic examination of a blood smear for malaria (3 tests) | 0.995 | 0.99 | 0.999 | B | (12, 13) |
| Polymerase chain reaction (PCR) test for malaria | 1 | 1 | 1 | NA | Assumption |
| **Test specificity** | | | | | |
| Two stool O&P tests for *Ascaris*, hookworm, or *Trichuris* | 1 | 1 | 1 | NA | (1) |
| *Schistosoma* serologic test (ELISA) | 0.84 | 0.79 | 0.88 | B | (10) |
| *S. stercoralis* serologic test (IVD ELISA) | 0.97 | 0.96 | 0.99 | B | (11) |
| Microscopic examination of a blood smear for malaria (3 tests) | 1 | 0.98 | 1 | NA | (12, 13) |
| PCR test for malaria | 1 | 1 | 1 | NA | Assumption |
| **Baseline infection prevalence (without overseas presumptive treatment)** | | | | | |
| *Ascaris* | 0.106 | 0.053 | 0.159 | B | (1, 14) |
| Hookworm | 0.018 | 0.009 | 0.027 | B | (1, 14) |
| *Trichuris* | 0.077 | 0.039 | 0.116 | B | (1, 14) |
| *Schistosoma* | 0.091 | 0.003 | 0.327 | B | (15) |
| *S. stercoralis* | 0.109 | 0.024 | 0.242 | B | (15) |
| *Plasmodium spp.* (malaria) | 0.072 | 0.045 | 0.1425 | B | (16-24) |
| **Duration of infection (year)** | | | | | |
| *Ascaris* | 1 | 1 | 2 | G | (25, 26) |
| Hookworm | 6 | 1 | 7 | G | (25, 27) |
| *Trichuris* | 2 | 1 | 2 | G | (25, 28) |
| *Schistosoma spp.* | 5 | 3 | 10 | G | (29) |
| *S. stercoralis* | lifetime | lifetime | lifetime | NA | Assumption, (30) |
| *Plasmodium falciparum* (malaria) ^b^ | 1 | 1 | 1 | NA | Assumption, (31) |
| **Annual probability of seeking treatment given infection** | | | | | |
| Outpatient visit for ascariasis/hookworm/ trichuriasis/schistosomiasis | 0.001 | 0.00012 | 0.005 | B | (1, 32, 33) |
| Outpatient visit for strongyloidiasis | 0.001 | 0.00012 | 0.005 | B | (1, 32-34) |
| Inpatient strongyloidiasis | 2.9E-05 | 6.6E-06 | 1.2E-04 | B | (1, 32-34) |
| Outpatient visit for malaria | 0.015 | 0.008 | 0.025 | B | (18, 35-39) |
| Inpatient malaria | 0.029 | 0.015 | 0.047 | B | (18, 35-39) |
| Case fatality rate (CFR) for inpatient strongyloidiasis | 0.167 | 0.085 | 0.6425 | B | (34) |
| CFR for inpatient malaria | 0.0072 | 0.0048 | 0.0154 | B | (36, 40) |

^a^ Distribution types: B- generalized beta, G- gamma, NA-Not available

^b^ We used one year because we assume that *Plasmodium falciparum* will be the most common infection among U.S.-bound refugees.{Debash, 2025 #248}

Drug efficacy

The U.S. Centers for Disease Control and Prevention (CDC) recommends using albendazole to treat *Ascaris*, hookworm, and *Trichuris* infections (41). The efficacy of albendazole against each infection was estimated from meta-analyses of cure rates, i.e., the presence or absence of helminth eggs in stool after treatment (4). World Health Organization (WHO) recommends the use of egg reduction rates, rather than cure rates, to be used as the critical indicator of drug efficacy against these infections (42). The use of cure rates would result in a more conservative estimate of efficacy in comparison to the use of egg reduction rates. Our analyses used cure rates, which would yield a conservative estimate. (Table S2).

Table S2: Estimates of cure rates and egg reduction rates against ascariasis (*Ascaris lumbricoides*), hookworm, and trichuriasis (*Trichuris trichuria)* for albendazole (4)

|  | Cure Rate | | Egg Reduction Rate | |
| --- | --- | --- | --- | --- |
|  | % | 95% CI | % | 95% CI |
| *A. lumbricoides* | 96.5 | 94.4-97.9 | 99.7 | 99.2-99.9 |
| Hookworm | 78.5 | 71.5-84.2 | 92.1 | 88.5-94.8 |
| *T. trichuria* | 32.1 | 23.2-42.5 | 64.3 | 53.2-73.5 |

Praziquantel is the recommended drug of choice for treating schistosomiasis (*Schistosoma* infections) (43). For S. mansoni, S. haematobium, and S. intercalatum infections, the U.S. CDC recommends administering 40 mg/kg orally in two divided doses over one day (43). Since our study population was limited to African refugees, we did not consider *S. japonicum* or *S. mekongi* infections, which are common in Asia. These infections would require a higher dose of 60 mg/kg orally in three divided doses over one day (43). The probabilities of parasitological failure with praziquantel 40 mg/kg, as reported in Cochrane Database Systematic Reviews, were 22% (range: 5% to 48%) for S. mansoni infections (5) and 38% (range: 26% to 54%) for S. haematobium infections (6) in endemic areas. We used the probability of parasitological success (i.e., 1– probability of parasitological failure for each) to estimate the efficacy of praziquantel against each *Schistosoma* species.

Ivermectin has been recommended for the treatment of *Strongyloides stercoralis* infections (44). According to a Cochrane Database Systematic Review, the overall cure rate of ivermectin against *S. stercoralis* infection was 84% (range: 72% to 98%) (7). The U.S. CDC recommends using the fixed combination of artemether-lumefantrine (AL; Coartem®) as a presumptive treatment for *P. falciparum* malaria for all U.S.-bound refugees from sub-Saharan Africa (45). The efficacy of AL (Coartem®) against *P. falciparum* malaria was 97% (range: 96% to 98%) from a recent systematic review (8).

Test sensitivity and specificity

The sensitivity and specificity of screening tests were estimated for each parasite. We assumed that two stool ova and parasite (O&P) tests would be performed for the diagnosis of *Ascaris*, hookworm, and *Trichuris* infections; two serologic tests would be performed for the diagnosis of *Schistosoma* (one test) and *S. stercoralis* (one test), and either microscopic examination of a blood smear (three repeated tests) or polymerase chain reaction (PCR) test would be performed for malaria diagnoses. For the screen and treat alternative, we assumed that all these tests would be conducted during the initial comprehensive medical exams in the absence of presumptive overseas treatment.

The sensitivity and specificity of tests for diagnoses of *Ascaris*, hookworm, and *Trichuris* infections were obtained from Nikolay et al. 2014 (9) and Maskery et al. 2016 (1) (Table S1). When repeated tests were used (i.e. for ascariasis, hookworm, trichuriasis, or malaria), the baseline estimate was estimated using the following equation: 1 – (1 – sensitivity of a single test)^x^ where *x* is the number of repeated tests. (Note that equation is simplified by the assumption of 100% specificity of repeated stool O & P tests or for malaria blood smears.) The estimate assumed that a positive test result would be recorded when either of the two samples tested positive for ascariasis, hookworm, or trichuriasis or one of the three samples for malaria. The sensitivity and specificity of the Enzyme-Linked Immunosorbent Assay (ELISA) test for the diagnosis of *Schistosoma* infections were obtained from Beltrame et al. 2017 (10). The sensitivity and specificity of in vitro diagnostic (IVD) ELISA tests for diagnoses of *S. stercoralis* infections were obtained from Bisoffi et al. 2014 (11). For malaria diagnoses, we assumed that the PCR test has 100% sensitivity and specificity. The sensitivity and specificity of microscopic examination of a blood smear for malaria diagnoses were obtained from Martín-Díaz et al. 2018 and Badiane et al. 2022 (12, 13).

Baseline infection prevalence (without overseas presumptive treatment)

The baseline prevalence of *Ascaris*, hookworm, and *Trichuris* infections among refugees from Africa without presumptive overseas treatment programs was estimated from a study of newly-arrived refugees (9) and summarized in Table S3. The prevalence data from African refugees who may have been treated presumptively with albendazole were adjusted using drug efficacy (4), and sensitivity and specificity of diagnostic tests (1). The following equation was used to estimate the baseline prevalence of the three soil-transmitted helminth infections among refugees from Africa without overseas presumptive treatment programs.

${Prevalence}_{u}=\frac{( {Prevalence}_{t}+Specificity-1)}{\left( Sensitivity+Specificity-1 \right)\times(1-efficacy)}$

$${Prevalence}_{u}:Prevalence before treatment$$

$${Prevalence}_{t}:Prevalence after treatment$$

We assumed a range of ±50% of the baseline values for the lower and upper bound estimates.

Table S3: Estimated prevalence of *Ascaris*, hookworm, and *Trichuris* infections

|  | *Ascaris* | hookworm | *Trichuris* |
| --- | --- | --- | --- |
| Prevalence (Treated) | 0.3% | 0.3% | 5.0% |
| Drug efficacy | 97% | 79% | 32% |
| Adjusted pre-treatment prevalence | 8.6% | 1.4% | 7.4% |
| Sensitivity of test | 81% | 78% | 96% |
| Specificity of test | 100% | 100% | 100% |
| Estimated baseline prevalence (Untreated) | 10.6% | 1.8% | 7.7% |

The prevalence of *Schistosoma* and *S. stercoralis* infections was estimated from a meta-analysis (15). Schistosomiasis seroprevalence among migrants from combined Africa, Middle East and North Africa, and sub-Saharan Africa were 9.1% [95% Confidence Intervals (CI): 2.8%-18.4%], 6.4% [95% CI: 0.3%-19.5%], and 24.1% [95% CI: 16.4%-32.7%], respectively (15). Strongyloidiasis seroprevalence among migrants from all Africa was 10.9% [95% CI: 5.2%-18.3%]. When subdividing the continent into 1) Middle East and North Africa and 2) sub-Saharan Africa, the respective prevalence estimates were 5.5% [95% CI: 2.4%-9.7%] and 14.6% [95% CI: 7.1%-24.2%] (15). For our analysis, the estimates of seroprevalence among migrants from all Africa were used as baseline estimates. The lower bound estimates were the lower bound of the 95% CI of seroprevalence among migrants from the Middle East and North Africa. The upper bound estimates were the upper bound of the 95% CI of seroprevalence among migrants from sub-Saharan Africa.

The prevalence of malaria was estimated from multiple studies investigating malaria prevalence among African migrants, as summarized in Table S4. The U.S. CDC’s recommendation of overseas (pre-departure) presumptive anti-malaria therapy for all non-pregnant sub-Saharan African refugees has been implemented since May 1999 (18). Thus, the symptomatic malaria prevalence in the United States among sub-Saharan African refugees arriving after 1999 would have been estimated after anti-malarial treatment that would have been received by most of these refugees, and the estimates ranged between 0% and 1.6% (18, 22). To estimate baseline malaria prevalence (i.e., without overseas presumptive treatment), we reviewed studies that investigated malaria prevalence among African refugees or migrants arriving in countries that had not implemented presumptive treatment programs or among U.S.-bound refugees prior to undergoing presumptive treatment. The range of malaria prevalence from these studies was found to be from 1.3% among sub-Saharan immigrants screened for infectious diseases in Spain from January 2009 to December 2012 (24), to 23% among U.S.-bound Congolese refugees living in the Kikuube district, Uganda, in 2018 (21). Since this upper bound estimate was an outlier and was reported for a population that was chosen due to a high incidence of malaria, we used the second highest estimate of 14.25% as the upper bound for this analysis. The baseline estimate was 7.2% of the malaria incidence rate among the immigrant population coming from sub-Saharan Africa and living in a country not endemic for malaria <1 years, which is the most representative of our study.

Table S4. Prevalence of malaria among African refugees from selected publications

| Source (Country) | Reported malaria prevalence ^a^ | Descriptions |
| --- | --- | --- |
| Phares et al. 2011 (USA) (22) | 0.13%-1.6% | Incidence rates of symptomatic malaria among newly arrived refugees between May 2007 and February 2008 were 1.6% among those who received sulfadoxine-pyrimethamine during the overseas, pre-departure presumptive treatment, 0.3% among those who received AL under partial supervision, and 0.13% among those who received AL under full supervision. |
| Serre Delcor et al. 2016 (Spain) (24) | 1.3% | Two cases of asymptomatic malaria (1.3%, 2/157) by *P. falciparum* were diagnosed among sub-Saharan immigrants screened for infectious diseases at International Health Center in Spain from January 2009 to December 2012. |
| Monge-Maillo et al. 2015 (Spain) (20) | 4.5% | Among 222 asymptomatic Sub-Saharan African immigrants in Spain who attended to a European Tropical Medicine Referral Center from 2000 to 2009, the prevalence of malaria was 4.5% (10/222). |
| Salas-Coronas et al. 2018 (Spain) (23) | 7.2% | Among sub-Saharan immigrants who newly arrived in Spain and attended the Tropical Medicine Unit of the Hospital de Poniente from October 2004 to February 2017, 7.2% of them (35/488) were positive for malaria with rapid diagnostic tests (RDT) confirmed by PCR tests. |
| Cobo et al. 2016 (Spain) (17) | 7.9% | The malaria incidence rate among immigrant populations coming from sub-Saharan Africa and living in Spain for <3 years was 7.9% (100/1,255), while the incidence rate among sub-Saharan immigrants living in Spain for >3 years was 1.0% (7/675) in the Tropical Medicine Unit of the Hospital of Poniente (Spain) from October 2004 to December 2013. |
| Collinet-Adler et al. 2007 (USA) (18) | 8.2% | During the pre-implementation stage (1996-1998) of the overseas, pre-departure treatment program, when the pre-departure treatment coverage rate was less than 10%, the symptomatic malaria incidence rate was 8.2% (27/330) among West African refugees in Hennepin County (Minneapolis), Minnesota. |
| Bocanegra et al. 2014 (Spain) (16) | 9.3% | Among sub-Saharan immigrants who attended the Tropical Medicine Unit of the Vall d’Hebron Teaching Hospital (Barcelona) in Spain between September 2007 and March 2010, 9.3% of them tested positive for malaria (25/267). |
| Corbacho-Loarte  et al. 2022 (Spain) (19) | 14.25% | From 2010-2019, 14.25% of asymptomatic sub-Saharan migrants in Spain tested positive for malaria (90/632) among patients attended at the National Reference Unit for Tropical Diseases in Spain. |
| Mwesigwa et al. 2021 (Uganda) (21) | 23% | During February-March 2018, 23% (187/803) of U.S.-bound Congolese refugees living in the Kikuube district, Uganda, were positive for malaria by either RDT or PCR. (Before malaria treatment) |

^a^ Most of these studies used a convenience sample of individuals referred to hospitals specializing in tropical medicine in which all referred patients migrated from sub-Saharan Africa. All such patients were screened for malaria. In contrast, the Collinet-Adler et al. 2022 and Phares et al. 2011 studies calculated incidence based on individuals who sought treatment for malaria (18, 22). The Mwesigwa et al. 2021 study screened all U.S.-bound refugees living in Kikuube district, Uganda (21). The incidence rates obtained from passively detected cases would be expected to be lower than the prevalence rates obtained from screening all individuals from given populations.

Duration of infection

*Ascaris*, hookworm, *Trichuris*, and *Schistosoma spp.* are unable to replicate within the human host. As a result, once the human host is infected, the concentration of parasites will decrease over time unless the patient is reinfected. Estimates of the duration of infection are available based on the estimated lifespan of adult worms. The durations of infection were estimated to be one year for *Ascaris* (range: 1 to 2 years) (25, 26), six years for hookworm (range: 1 to 7 years) (25, 27), two years for *Trichuris* (range: 1 to 2 years) (25, 28), and five years for *Schistosoma* (range: 3-10 years) (29)*. S. stercoralis* is distinguished from other helminth infections due to its unique auto-infective life cycle, which can result in lifelong infection in the host (30). We assumed that the duration of infection for S. *stercoralis* was the lifetime of the host (i.e., this model considers 65 years of life after the infection).

It was previously believed that *P. falciparum* infections lasted for less than two years (31). However, some recent studies have shown evidence that asymptomatic *P. falciparum* infections may continue for up to ten years or longer (31). Despite these longer observed durations in subsets of the population, we decided to use an average duration of infection of one year since this was the period used to estimate the annual probability of seeking treatment given infection as summarized in the next section. This would lead to an underestimate of any disutility among individuals with asymptomatic or subclinical malaria infections.

Annual probability of seeking treatment given infection

Parasitic infections often exhibit an asymptomatic or subclinical course, particularly in individuals who have resided in endemic areas. As a result, a significant portion of patients with parasitic infections may go unnoticed, necessitating the estimation of the number of asymptomatic cases. This information serves as the denominator for calculating the annual probabilities of seeking treatment.

To calculate the annual probability of seeking malaria treatment among African refugees, we used multiple data sources. First, we reviewed 2018 malaria surveillance data in the United States to identify the number of African refugees or immigrants who reported inpatient or outpatient visits associated with malaria infections (36). In 2018, there were 375 reported cases of imported malaria cases among non-US residents, with 40 of them (10.7%) classified as severe malaria (36). Among the 375 non-US residents with reported cases of imported malaria, 143 patients were identified as refugees or immigrants from Africa (36). We assumed that 15 of them had severe malaria (143 × 10.7%), while the remaining 128 had uncomplicated malaria. Overall in 2018, 62.8% of those with uncomplicated malaria were hospitalized, and 93.6% of those with severe malaria were hospitalized (36). Based on these percentages, we estimated that 94 (15 × 93.6% + 128 × 62.8%) out of the 143 patients from Africa with immigrant or refugee status were hospitalized. We assumed that the remaining 49 malaria patients were not hospitalized and had outpatient visits only.

Next, we used the 2022 Yearbook of Immigration Statistics (39) and the malaria infection prevalence with and without overseas presumptive treatment. Using these data, we estimated the total number of individuals with malaria infection, including those who were asymptomatic, among African refugees or immigrants (Table S5). We estimated the number of refugees who arrived in the U.S. in 2018 by departing country from the 2018 Yearbook of Immigration Statistics and calculated the numbers of refugees from malaria-endemic areas in Africa (10,462) (39). Due to the implementation of presumptive treatment with AL for U.S.-bound refugees prior to their departure to the United States, we estimated that the baseline prevalence of malaria among these refugees who received presumptive treatment was 0.22%. This estimate was derived by considering the malaria prevalence without presumptive treatment (7.2%) and the efficacy (8) of AL against malaria (97%), calculated as 7.2% × (100% ‒ 97%). To estimate the lower bound estimate, we used the lower bound malaria prevalence (1.3%) without presumptive treatment and the upper bound efficacy of AL (8) against malaria (98%), resulting in an estimate of 0.03%. Conversely, the upper bound estimate (0.92%) was obtained by using the upper bound malaria prevalence (23%) without presumptive treatment and the lower bound efficacy (8) of AL against malaria (96%). We estimated the number of African refugees with malaria infection to be 23 (10,462 × 0.22%; range:3-96).

The number of immigrants from malaria-endemic areas in Africa was estimated to be 95,119 (35, 39). We assumed that the baseline prevalence of malaria among these immigrants may be lower than the estimated prevalence for refugees or irregular migrants, which comprised most of the estimates reported in Table S4. We could not identify studies that specifically compared immigrants and refugees with malaria infection. However, we did review a study that directly compared the rates of tuberculosis diagnosed in U.S.-bound immigrants compared to refugees. In the study, the rate of tuberculosis in immigrants was about 46% as high as that reported for refugees (220 per 100,000 for immigrants compared to 470 per 100,000 in refugees) (46). The resulting multiplier using tuberculosis as a proxy to estimate the potential difference in malaria prevalence is approximately 0.466. This multiplier was then applied to the prevalence estimate for refugees to estimate the prevalence in immigrants: 7.2% (range: 1.3%-23%), equivalent to the prevalence estimates from African refugees without treatment (Table S1). The estimated baseline number of immigrants from Africa with malaria infection, including asymptomatic individuals, was 3,192 (95,119 × 3.4%; range: 576-10,197).

Table S5: Estimated number of symptomatic and asymptomatic malaria patients among African refugees or immigrants arrived at the United States in 2018

|  | Baseline | Lower bound | Upper bound |
| --- | --- | --- | --- |
| Refugees | | | |
| Malaria presumptive treatment before the U.S. arrival | Yes | | |
| Number of arrivals in 2018, (A) | 10,462 | | |
| Prevalence of malaria, (B) | 0.22% | 0.03% | 0.92% |
| Estimated number of refugees with malaria infection, symptomatic and asymptomatic, (C)=(A) x (B) | 23 | 3 | 96 |
| Immigrants | | | |
| Malaria presumptive treatment before the U.S. arrival | No | | |
| Number of arrivals in 2018, (D) | 95,119 | | |
| Prevalence of malaria, (E) | 7.2% | 1.3% | 23% |
| Multiplier to estimate prevalence malaria in immigrants, (F1) | 0.466 | | |
| Estimated prevalence of malaria in immigrants (F2) | 3.36% | 0.61% | 10.72% |
| Estimated number of immigrants with malaria infection, symptomatic and asymptomatic, (G)=(D) × (F2) | 3,192 | 576 | 10,197 |
| Total number of individuals with malaria infection (refugees and immigrants), (C)+(G) | 3,215 | 579 | 10,293 |

Thus, the total number of individuals with malaria infection, symptomatic and asymptomatic, among refugees or immigrants who arrived in the United States in 2018 was 3,215 (range: 579-10,293) (Table S5). The estimated baseline annual probability of seeking malaria treatment among them was 2.9% (94÷3,215, range: 0.0%-16.2%) for inpatient visits and 1.5% (49÷3,215; range: 0.5%-8.5%) for outpatient visits.

The annual probabilities of seeking outpatient treatment for *Ascaris*, hookworm, *Trichuris*, *Schistosoma*, and *S. stercoralis*, as well as the annual probability of seeking inpatient treatment for strongyloidiasis were obtained from a published article (1), which incorporated estimates from earlier analyses (32-34).

The case fatality rates (CFR) were estimated specifically for hospitalized patients. We assumed that hospitalizations among patients with *Ascaris*, hookworm, *Trichuris*, and *Schistosoma* infections were negligible and thus focused on the CFRs for strongyloidiasis and malaria inpatients only. The CFR for strongyloidiasis inpatients was taken from a published article (34), while the CFR for malaria inpatients was estimated using the U.S. CDC malaria surveillance data from 2010 to 2018 (36, 40). We assumed that all malaria-related deaths were observed among malaria patients who were reported to be hospitalized. This may slightly overestimate the CFR because hospitalization status was not available for all identified malaria patients. The baseline estimate (0.0072) was calculated from the average CFR between 2010 and 2018, as shown in Table S6. The upper bound estimate was the maximum annual CFR of malaria inpatients between 2010 and 2018 (0.0154 in 2015). The lower bound estimate was the minimum value recorded during the same period (0.0048 in 2011).

Table S6: Estimated case fatality rate (CFR) among hospitalized malaria patient in the United States, 2010-2018 (36, 40)

| Year | Reported numbers of malaria hospitalization | Number of deaths associated with malaria | CFR among hospitalized malaria patient |
| --- | --- | --- | --- |
| 2010 | 946 | 9 | 0.0095 |
| 2011 | 1,047 | 5 | 0.0048 |
| 2012 | 1,023 | 6 | 0.0059 |
| 2013 | 1,003 | 10 | 0.0100 |
| 2014 | 766 | 5 | 0.0065 |
| 2015 | 714 | 11 | 0.0154 |
| 2016 | 1,261 | 7 | 0.0056 |
| 2017 | 1,302 | 7 | 0.0054 |
| 2018 | 1,223 | 7 | 0.0057 |
| Total | 9,285 | 67 | 0.0072 |

### Program parameters

The basic program parameters include whether presumptive treatment may be provided in all settings prior to refugee departure for the United States. In addition, pregnant individuals would be ineligible to receive presumptive treatment. As a result, the federal government would likely be unable to achieve 100% coverage for the presumptive treatment program. To account for these limitations in coverage, we assumed that between 80% and 100% (90% was used as the baseline) of individuals would be covered by presumptive treatment programs. Similarly, we assumed that between 80% and 100% (baseline: 90%) would be able to undergo a comprehensive medical examination after arrival in the United States. For individuals who are unable to receive presumptive treatment prior to departure, we assumed they may be tested and treated (if necessary) at their follow-up comprehensive examinations. Further, we assumed that individuals who received presumptive treatment would not be evaluated for these parasitic diseases at their comprehensive examinations. In contrast, we assumed “Domestic Screening and Treatment” and “Domestic Presumptive Treatment” would only provide one opportunity to be evaluated or treated. For simplification and to provide a more straightforward comparison, we assumed all individuals would undergo either screening and treatment or presumptive treatment for the domestic-only alternatives. For drug efficacies, we assumed that overseas treatment may be lower than domestic treatment due to the potential lower quality overseas drugs; thus, we added a correction factor (0.95, ranging between 0.9 and 1) to account for this.

Table S7: Summary of parameters associated with overseas presumptive treatment and domestic screening and treatment programs

| Description | Baseline | Lower bound | Upper bound | Distri-bution ^a^ | Ref |
| --- | --- | --- | --- | --- | --- |
| Proportion of refugees receiving domestic comprehensive medical exam | 0.9 | 0.8 | 1 | U | Assumptions |
| Probability refugees arrive from countries with presumptive treatment program and received presumptive treatment | 0.9 | 0.8 | 1 | U |  |
| Adjustment factor for overseas versus domestic treatment efficacy | 0.95 | 0.9 | 1 | U |  |

^a^ Distribution types: U- uniform

### Demographics

The background death rate varies depending on age. We used crude death rates per 100,000 persons by five-year age group from the underlying cause of death 2018-2021 data (Table S8) (47). The proportion of refugee arrivals by age was estimated using data from the Yearbook of Immigration Statistics for the years 2013-2022 (Table S8) (39). The average number of refugee arrivals from Africa was 15,656 and their mean age of refugee arrivals during this period was 25 years (39). The proportion of female refugee arrivals between 2012 and 2021 was 49.2% (39). We assumed that female refugees between the ages of 12 and 45 years old must undergo pregnancy tests, with an estimated proportion of refugees requiring pregnancy tests being 28.8%.

Table S8: Background death rate and proportion of refugee arrivals by age

| Age (year) | Death rate per 100,000 persons (47) | | Proportion of refugee arrivals by age (%) (39) |
| --- | --- | --- | --- |
|  | N | 95% CI |  |
| <1 | 548.5 | 544.7 − 552.2 | 0.3 |
| 1-4 | 23.7 | 23.3 − 24.1 | 10.2 |
| 5-14 | 13.7 | 13.5 − 13.9 | 22.4 |
| 15-24 | 78.3 | 77.8 – 78.7 | 20.5 |
| 25-34 | 149.4 | 148.9 – 150.0 | 20.1 |
| 35-44 | 233.1 | 232.4 – 233.9 | 12.6 |
| 45-54 | 447.8 | 446.8 − 448.8 | 7.1 |
| 55-64 | 981.9 | 980.4 − 983.4 | 4.0 |
| 65-74 | 1,948.7 | 1,946.3 – 1,951.2 | 2.1 |
| 75-84 | 4,709.3 | 4,704.0 – 4,714.6 | 0.8 |
| 85+ | 14,379.7 | 14,365.1 – 14,394.3 |  |
| Total | 952.2 | 951.9 – 953.0 | 100 |

### Economic input parameters estimation

The economic input parameters are summarized in Table S9.

Table S9. Summary of economic input parameters (2021 USD)

| Description | Baseline | Lower bound | | Upper bound | | Distri-bution ^a^ | | Ref | |
| --- | --- | --- | --- | --- | --- | --- | --- | --- | --- |
| **Domestic screening for intestinal parasites and malaria** | | | | | | | | | |
| Comprehensive exam | 15 | 14 | | 16 | | G | | (48, 49) MarketScan | |
| Stool ova and parasites tests | 17 | 16 | | 18 | | G | |  |  |
| Helminth antibody tests | 33 | 25 | | 38 | | G | |  |  |
| Malaria screening tests | 28 | 20 | | 32 | | G | |  |  |
| Translator | 9 | 9 | | 9 | | NA | | (50) | |
| Total costs for domestic screening | 102 | 85 | | 113 | | G | |  | |
| **Treatment costs for refugees diagnosed with intestinal parasites or malaria infections during comprehensive exams in the United States after arrival** | | | | | | | | | |
| Hookworm or *Ascaris* infections | 185 | 84 | | 633 | | G | | (41, 51, 52) | |
| *Trichuris* infections | 458 | 150 | | 1,798 | | G | | (41, 51, 52) | |
| *Schistosoma* infections | 236 | 234 | | 350 | | G | | (43, 51, 53) | |
| *S. stercoralis* infections | 70 | 67 | | 84 | | G | | (44, 51, 54) | |
| Malaria infections | 172 | 170 | | 198 | | G | | (51, 55, 56) | |
| **Opportunity costs for domestic screening** | | | | | | | | | |
| Domestic screening for intestinal parasites and malaria | 10 | 10 | | 34 | | G | | (50, 57) Assumptions | |
| Follow visit after intestinal parasites or malaria diagnosis | 8 | 8 | | 28 | | G | |  |  |
| **Outpatient treatment costs without an intervention** | | | | | | | | | |
| *Ascaris* infections | 340 | 164 | | 863 | | G | | (41, 51, 52, 58, 59) | |
| Hookworm infections | 391 | 262 | | 866 | | G | |  |  |
| *Trichuris* infections | 795 | 424 | | 2,198 | | G | | (41, 51, 52, 58, 59) | |
| *Schistosoma* infections | 848 | 525 | | 1,138 | | G | | (43, 51, 53, 58, 60) | |
| *S. stercoralis* infections | 577 | 449 | | 717 | | G | | (44, 51, 54) MarketScan | |
| Malaria infections | 911 | 581 | | 1,240 | | G | | (58, 61) | |
| **Inpatient treatment costs without an intervention** | | | | | | | | | |
| *S. stercoralis* infections | 181,359 | 21,678 | | 303,926 | | G | | (58) MarketScan | |
| Malaria infections | 20,405 | 8,708 | | 32,101 | | G | | (58, 61) | |
| **Opportunity costs for outpatient and inpatient visits** | | | | | | | | | |
| Outpatient | 192 | 192 | | 224 | | G | | (50, 57)  Assumption | |
| Strongyloidiasis inpatient | 2,496 | 768 | | 3,360 | | G | | (50, 57) MarketScan | |
| Malaria inpatient | 1,536 | 960 | | 1,792 | | G | | (58, 61) Assumption | |
| **Overseas cost estimates** | | | | | | | | | |
| Presumptive albendazole, praziquantel, ivermectin, and AL treatment in Africa | 29 | | 22 | | 37 | | G | | 2023 IOM data |

^a^ Distribution types: G- gamma

#### Costs of domestic screening and treatment for intestinal parasite and malaria

Each refugee was assumed to undergo a comprehensive exam to assess several health concerns, including parasitic infections. The cost for domestic screening of intestinal parasites and malaria would comprise a fraction of the comprehensive exam costs for new patients (Current Procedural Terminology (CPT^®^) code: 99204), costs for diagnostics (such as two stool O&P tests (CPT^®^ code: 87177) for *Ascaris,* hookworm, and *Trichuris*, two helminth antibody tests (CPT^®^ code: 86682) for *S. stercoralis* and *Schistosoma*, and three microscopic examinations of blood smears (CPT^®^ code: 87207) or molecular diagnosis using PCR (CPT^®^ code: 87798) for malaria), and costs for translators. For the cost of malaria screening, a weighted average of 80% of the blood smear cost and 20% of the PCR cost was used, assuming that a blood smear is the most used test for malaria screening and that three blood smear tests would be performed. We assumed that 10% of the cost of the comprehensive exam could be attributable to screening for intestinal parasites and malaria, and the remaining 90% could be used for other health conditions in refugees.

Cost estimates were based on payments for patients with public or private health insurance. The total payments (insurance + patient co-payments) for individuals with public health insurance were estimated using the 2021 Medicare Clinical Laboratory Fee Schedule or the facility prices of the 2021 Medicare Physician Fee Schedule (MPFS) from the Centers for Medicare and Medicaid Services (CMS) (48, 49). Payments for patients with private insurance were estimated using the Merative^®^ MarketScan^®^ Commercial Database (Merative^®^, Ann Arbor, MI) using the 10 million subsample Set B from January 1, 2021, to December 31, 2021, with an online tool called MarketScan^®^ Treatment Pathways. We excluded the following patients:

(1) Patients with capitated health plans,

(2) Patients who reported zero payments associated with the selected CPT^®^ codes, and

(3) Patients with payment outliers by applying the interquartile range (IQR) method.

Values that were more than 1.5 IQR below the first quartile or more than 1.5 IQR above the third quartile were considered outliers (62). The baseline estimate was the mean total payment associated with each CPT^®^ code. We used the first and third-quartile numbers for the lower and upper-bound estimates.

The cost of domestic screening was estimated using a weighted average approach, incorporating 25% of MarketScan^®^ Commercial Database payments and 75% of CMS reimbursement rates (Table S10). Baseline estimates were obtained as a weighted average of mean payments from the MarketScan^®^ Commercial Database and the CMS reimbursement rates. Upper and lower bound estimates were derived from the upper and lower bound estimates from MarketScan® Commercial Database and the CMS reimbursement rates. These weights were assumed because we think that most refugees would have access to Medicaid upon arrival (as opposed to private insurance).

Additionally, we assumed that all refugees from Africa would need language translation assistance during the initial comprehensive exam, with 15 minutes (i.e., 0.25 hours) of the translator’s time dedicated to intestinal parasites and malaria. The cost of the translator was estimated based on the mean hourly wage for translators (occupation code 27-3091, $28.08) from May 2021 National Occupational Employment and Wage Estimates, U.S. Bureau of Labor Statistics (BLS) (50). In addition, we accounted for non-wage benefits by adding 33% to the average hourly wage estimate. Thus, the translation cost was estimated from 0.25 hours × $28.08 hourly wage × 1.33 adjustment for non-wage benefits = $9.34.

Table S10. Costs for domestic screening for intestinal parasites and malaria in the absence of overseas presumptive treatment (2021 USD)

|  |  | CPT code | No. tests or visits | Baseline (USD) | Lower bound (USD) | Upper bound (USD) |
| --- | --- | --- | --- | --- | --- | --- |
| Comprehensive exam (new patient, level 4 complexity) | MarketScan^®^ | 99204 | 0.1^a^ | 189 | 156 | 219 |
|  | CMS |  |  | 137 | 137 | 137 |
|  | Weighted  (25% × MarketScan^®^ + 75% × CMS) |  |  | 150 | 142 | 158 |
| Stool ova and parasites test (*Ascaris*, hookworm, and *Trichuris*) | MarketScan^®^ | 87177 | 2 | 8 | 7 | 9 |
|  | CMS |  |  | 9 | 9 | 9 |
|  | Weighted  (25% × MarketScan^®^ + 75% × CMS) |  |  | 9 | 8 | 9 |
| Helminth antibody tests  (*S. stercoralis* and *Schistosoma*) | MarketScan^®^ | 86682 | 2 | 26 | 11 | 37 |
|  | CMS |  |  | 13 | 13 | 13 |
|  | Weighted  (25% × MarketScan^®^ + 75% × CMS) |  |  | 16 | 12 | 19 |
| Malaria screening tests | Weighted (80% × blood smear test + 20% × PCR) |  |  | 28 | 20 | 32 |
| a. Thick/thin blood smear test | MarketScan^®^ | 87207 | 3 | 45 | 15 | 60 |
|  | CMS |  |  | 18 | 18 | 18 |
|  | Weighted  (25% × MarketScan^®^ + 75% × CMS) |  |  | 25 | 17 | 29 |
| b. Polymerase chain reaction (PCR) test | MarketScan^®^ | 87798 | 1 | 59 | 28 | 71 |
|  | CMS |  |  | 35 | 35 | 35 |
|  | Weighted  (25% × MarketScan^®^ + 75% × CMS) |  |  | 41 | 33 | 44 |
| Translator (0.25 hours × $37.35/hour) | NA | NA | 1 | 9 | 9 | 9 |
| Total |  |  |  | 102 | 85 | 113 |

^a^ This is based on the assumption that 10% of exam time would be devoted to screening for parasitic diseases.

Patients diagnosed with intestinal parasites or malaria during their comprehensive exams were assumed to return for a follow-up outpatient visit (complexity level 2) and receive medications for treatment. The costs associated with these treatments are summarized in Table S11. Baseline and lower bound estimates for drug costs per pill were based on Medicaid data for 2021 (51), with the exception of albendazole. The lower bound costs for albendazole were obtained from the Mark Cuban Cost Plus Drug Company website (63) because the reported price was lower than the Medicaid cost. Upper bound estimates were obtained from the Merative Micromedex^®^ RED BOOK^®^ database for brand-name drugs (Albenza^®^ for albendazole, Stromectol^®^ for ivermectin, Biltricide^®^ for praziquantel, and Coartem^®^ for a combination of 20 mg artemether and 120 mg lumefantrine) in 2021 (52-54, 56). We assumed that treatment would require the following (45):

(1) 2 × 200 mg albendazole for hookworm or *Ascaris* infections (41),
(2) 6 × 200 mg albendazole (400 mg orally for 3 days) for *Trichuris* infections (41),
(3) 3 × 600 mg praziquantel for *Schistosoma* infections (43),
(4) 6 × 3 mg ivermectin for *S. stercoralis* infections (44), or
(5) 6 doses × 4 tablets containing 20 mg artemether and 120 mg lumefantrine (Coartem®) per dose (three-day course with an initial dose, second dose after 8 hours, and then twice-daily for the following two days) for malaria (55).

We assumed that the above medications would be required and used the dosages from the overseas presumptive treatment program, which were also summarized in Table S18, except medication requirement for *Trichuris* infections. We assumed that most African refugees would not be exposed to *Schistosoma japonicum,* or *Schistosoma mekongi*.

Table S11. Treatment costs for refugees diagnosed with intestinal parasites or malaria infections during comprehensive exams in the United States after arrival (2021 USD)

|  | | Baseline (USD) | Lower bound (USD) | Upper bound (USD) |
| --- | --- | --- | --- | --- |
| Outpatient visit (existing patient, level 2 complexity, CPT code 99212) | MarketScan^®^ | 49 | 39 | 57 |
|  | CMS | 36 | 36 | 36 |
|  | Weighted  (25% × MarketScan^®^ + 75% × CMS) | 39 | 37 | 42 |
| Translator (0.25 hour $\times$ $37.35/hour) | | 9 | 9 | 9 |
| Albendazole (2 × 200mg) for *Ascaris* or hookworm | | 136 | 38 | 582 |
| Albendazole (3 days × 2 × 200mg) for *Trichuris* | | 409 | 104 | 1,747 |
| Praziquantel (3 × 600mg) for *Schistosoma* | | 187 | 187 | 299 |
| Ivermectin (2 days × 3 × 3mg) for *S. stercoralis* | | 21 | 21 | 33 |
| Coartem^®^ (6 doses × 4 tablets, Tablet=20mg artemether/ 120 mg lumefantrine) for malaria | | 123 | 123 | 147 |
| **Total treatment cost estimates ^a^** | | | | |
| Total for hookworm or *Ascaris* infections | | 185 | 84 | 633 |
| Total for *Trichuris* infections | | 458 | 150 | 1,798 |
| Total for *Schistosoma* infections | | 236 | 234 | 350 |
| Total for *S. stercoralis* infections | | 70 | 67 | 84 |
| Total for malaria infections | | 172 | 170 | 198 |

^a^ All estimated costs were calculated assuming that none of the refugees would have co-infections with multiple parasitic diseases. This assumption was made to simplify the analysis. The total treatment costs included costs of outpatient visits, translators, and medications for each infection.

Opportunity costs for domestic screening

The opportunity costs associated with domestic screening for African refugees were estimated based on assumed time requirements for undergoing comprehensive exams, providing stool samples, and making follow-up outpatient visits after diagnoses (Table S12). We assumed that two hours would be required for the comprehensive exam, including travel to and from appointments. The time required for comprehensive exams was prorated, as discussed above. Additionally, we assumed that one hour would be needed for providing stool samples (0.5 hrs. for two trips) and an additional hour to return for a follow-up outpatient visit after diagnosis. The value of time was estimated using two sources: 1) the baseline and lower bound cost estimates were based on the average U.S. Gross Domestic Product (GDP) per capita-hour using 2021 World Bank data ($70,249 per year $\div$ 365 days per year $\div$ 24 hours per day = $8 per hour) (57), and 2) the upper bound cost estimate was based on the average hourly wage estimate across all occupations from the 2021 BLS ($28 per hour) (50). We considered the average GDP per capita-hour as the baseline cost estimate because most refugees may not be employed during the initial comprehensive exam or may be able to undertake the exam during non-working hours.

Table S12. Opportunity cost estimates for African refugees participating in domestic screening programs (2021 USD)

|  | Time required (hours) | No. of tests or proportions of visits | Average GDP per capita-hour (USD/hour) | Average wage per hour (USD/hour) | Baseline and lower bound estimate (USD) | Upper bound estimate (USD) |
| --- | --- | --- | --- | --- | --- | --- |
| **Domestic screening for intestinal parasites and malaria** | | | | | | |
| Comprehensive exam | 2 | 0.1 | 8 | 28 | 2 | 6 |
| Time to provide two stool samples | 0.5 | 2 | 8 | 28 | 8 | 28 |
| Total opportunity cost | NA | | | | 10 | 34 |
| **Time required for follow-up visit after intestinal parasites or malaria diagnosis** | | | | | | |
| Outpatient visit | 1 | 1 | 8 | 28 | 8 | 28 |

#### Outpatient and inpatient treatment cost estimates

Outpatient treatment costs associated with intestinal parasites or malaria included outpatient visit costs and drug costs (Table S13). The outpatient visit costs for treating *Ascaris*, hookworm, and *Trichuris* were estimated using values from Joo et al. 2021 (59). The lower bound estimate was the mean outpatient visit costs for patients with Medicaid, while the upper bound estimate was based on the mean outpatient visit costs for commercially insured patients. The baseline estimate was calculated as the mean of the lower and upper bound estimates.

For treating *Schistosoma* infections, the baseline estimate was based on the mean of outpatient visit costs from patients with commercial insurance in 2015-2018 (60), with the lower and upper bound estimates derived from the lower and upper bound of the 95% CIs. Outpatient visit costs for treating *S. stercoralis* were estimated using the Merative® MarketScan® Commercial Database (Merative®, Ann Arbor, MI) 10 million subsample Set B from January 1, 2015, to December 31, 2020, with an online tool MarketScan® Treatment Pathways. The baseline estimate was the mean of outpatient visit costs, with the lower and upper bound estimates derived from the lower and upper bound of the 95% CIs. All estimated outpatient visit costs were adjusted to 2021 USD using the U.S. Medical Care Consumer Price Index (CPI) (58). Outpatient drug costs for intestinal parasites are from Table S11.

The estimated outpatient cost for malaria treatment was based on reported costs in Park et al. 2023 (61). As Park et al. 2023 did not separately report drug payments for Medicaid, we used the overall estimates of outpatient costs (including drug costs) for malaria from October 2015 to December 2019 (51). We used the same approach that was used to estimate the baseline, lower and upper bounds for the outpatient visit costs for treating *Ascaris*, hookworm, and *Trichuris,* as described above, in estimating the total outpatient costs for malaria. All costs were adjusted to 2021 USD using the U.S. Medical Care CPI (58).

Table S13: Outpatient visit and drug costs (2021 USD)

|  | Baseline (USD) | Lower bound (USD) | Upper bound (USD) |
| --- | --- | --- | --- |
| **Outpatient visit cost** | | | |
| *Ascaris* infections | 203 | 126 | 281 |
| Hookworm infections | 254 | 224 | 284 |
| *Trichuris* infections | 385 | 320 | 451 |
| *Schistosoma* infections | 661 | 338 | 839 |
| *S. stercoralis* infections | 556 | 428 | 684 |
| Malaria infections | 782 | 558 | 1,093 |
| **Outpatient drug cost** | | | |
| Albendazole (2 × 200mg) for hookworm or *Ascaris* | 136 | 38 | 582 |
| Albendazole (3 days × 2 × 200mg) for *Trichuris* | 409 | 104 | 1,747 |
| Praziquantel (3 × 600mg) for *Schistosoma* | 187 | 187 | 299 |
| Ivermectin (2 days × 3 × 3mg) for *S. stercoralis* | 21 | 21 | 33 |
| Coartem® (6 doses × 4 tablets, Tablet=20mg artemether/ 120 mg lumefantrine) for malaria | 123 | 123 | 147 |
| **Total** | | | |
| *Ascaris* infections | 340 | 164 | 863 |
| Hookworm infections | 391 | 262 | 866 |
| *Trichuris* infections | 795 | 424 | 2,198 |
| *Schistosoma* infections | 848 | 525 | 1,138 |
| *S. stercoralis* infections | 577 | 449 | 717 |
| Malaria infections | 911 | 581 | 1,240 |

Hospitalization costs associated with strongyloidiasis or malaria were obtained from the literature (Table S14) (34, 61). All estimated costs were adjusted to 2021 USD using the U.S. Medical Care CPI (58).

Table S14: Hospitalization costs associated with strongyloidiasis or malaria (2021 USD)

|  | Baseline (USD) | Lower bound (USD) | Upper bound (USD) |
| --- | --- | --- | --- |
| *S. stercoralis* infections | 181,359 | 21,678 | 303,926 |
| Malaria infections | 20,405 | 8,708 | 32,101 |

Opportunity costs for outpatient and inpatient visits

Although we do not use opportunity costs to calculate our incremental cost effectiveness ratios, we do report opportunity costs separately. Patient opportunity costs were estimated by considering the value of time and the loss of days associated with outpatient or inpatient visits (Table S15). The value of time was estimated using two sources: 1) the baseline and lower bound cost estimates were based on the average U.S. GDP per capita-hour ($192 per day) (57), and 2) the upper bound cost estimate was based on the average hourly wage estimate across all occupations from the 2021 BLS ($224 per workday) (50). We assumed that outpatient treatment would be associated with losing one full day for intestinal parasites or malaria.

Table S15: Estimated opportunity costs for outpatient visits associated with intestinal parasites or malaria

| Time required (days) | Average GDP per capita hour (USD/day) | Average wage per hour (USD/workday) | Baseline/ Lower bound estimate (USD) | Upper bound estimate (USD) |
| --- | --- | --- | --- | --- |
| 1 | 192 | 224 | 192 | 224 |

We estimated the opportunity costs for hospitalization based on the duration of inpatient treatment for strongyloidiasis or malaria patients (Table S16). We assumed that all other intestinal parasite infections, except for strongyloidiasis, would not result in hospitalization. The length of stay for strongyloidiasis inpatients was estimated using the Merative® MarketScan® Commercial Database (Merative®, Ann Arbor, MI) 10 million subsample Set B from January 1, 2014, to December 31, 2020, with an online tool MarketScan® Treatment Pathways. We excluded patients with capitated health plans and outliers using the IQR method described above. The baseline estimate for length of stay was set at the mean for hospitalized patients with strongyloidiasis (14 days). We used the first quartile (4 days) and third quartile (19 days) from this sample for the lower and upper bound estimates, respectively. For malaria hospitalizations, the lower and upper bound estimates for length of stays were 5 days and 10 days for patients with private insurance and Medicaid, respectively (61). The baseline estimate for malaria hospitalizations was the rounded average of the lower and upper bound estimates (8 days). We used the same methodology to account for the value of patient time as was described for outpatient visits above. We assumed 5 workdays per week to estimate the upper bound opportunity cost estimates, which incorporated the average wage to value time lost (i.e., 15 workdays for *S. stercoralis* infections and 8 workdays for malaria).

Table S16: Estimated opportunity costs for hospitalizations associated with strongyloidiasis or malaria

|  | Baseline | Lower bound | Upper bound |
| --- | --- | --- | --- |
| Hospitalizations associated with *S. stercoralis* infections | | | |
| Length of stay (days) | 13 | 4 | 19 |
| Opportunity cost per day (USD per day) | 192 | 192 | 224 |
| Estimated opportunity cost for *S. stercoralis* infections inpatient (USD) | 2,496 | 768 | 3,360 |
| Hospitalizations associated with malaria | | | |
| Length of stay (days) | 8 | 5 | 10 |
| Opportunity cost per day (USD per day) | 192 | 192 | 224 |
| Estimated opportunity cost for malaria inpatient (USD) | 1,536 | 960 | 1,792 |

### Overseas presumptive treatment cost

International Organization for Migration (IOM) has provided the budget for the fiscal year (FY) 2023 regarding the presumptive treatment program in Africa. The cost estimates included the following:

1. Staff cost.
2. Operational costs, which include costs for drug and pregnancy tests.
3. IOM overhead costs.

The IOM’s budget for FY2023 was formulated based on a projection of 5,000 U.S.-bound refugees from Africa.

The IOM staffing costs included salaries for one local staff member who allocated 25% of his/her time to the program for data entry, and two local IOM nurses who dedicated 100% of their time to drug preparation, pregnancy tests, and monitoring additional drug doses (Table S17). Their monthly salaries were $2,000 and $2,500 per person, respectively. The annual costs for these staff totaled $66,000including data entry ($2,000 per month × 25% × 12 months) and two IOM nurses ($2,500 per month × 100% × 12 months). An additional 30% of this staff cost ($66,000 × 30%) was allocated as shared costs for IOM administration or supporting staff. Consequently, the total annual staff cost for the overseas presumptive treatment program for African refugees was $85,800. The annual staff cost per African refugee was $17.16 ($85,800/5,000).

Table S17: Staff costs for overseas presumptive treatment costs for U.S.-bound refugees in Africa

| Staff Type | Number of staff, (a) | Monthly salary per person, (b) | Proportion of staff’s time for presumptive treatment, (c) | Annual cost, (d)=(a)×(b)×12 months×(c) |
| --- | --- | --- | --- | --- |
| Local data entry staff, (e) | 1 | 2,000 | 25% | $6,000 |
| Local IOM nurse, (f) | 2 | 2,500 | 100% | $60,000 |
| Administration / Support staff, (g) = ((e)+(f))×30% | NA | NA | NA | $19,800 |
| Total annual staff cost, (h)=(e)+(f)+(g) | NA | NA | NA | $85,800 |

Operational costs encompassed drug costs (albendazole, praziquantel, ivermectin, and AL) as well as the costs of pregnancy tests for female refugees between 12 and 45 years old. The estimated drug costs are summarized in Table S18. The average costs per person for each drug were as follows: $0.10 for albendazole, $1.00 for praziquantel, $6.00 for ivermectin, and $3.00 for AL. Consequently, the average total drug costs per person amounted to $10.1.

Table S18: Drug costs for overseas presumptive treatment for U.S.-bound refugees in Africa

| Drug | Average unit cost | Average dosage per person | Average drug costs per person |
| --- | --- | --- | --- |
| Albendazole | $0.10 per 400 mg (1 tablet) | 400 mg per person | $0.10 |
| Praziquantel | $1.00 per 1800 mg (3 tablets) | 1800 mg per person | $1.00 |
| Ivermectin | $2.00 per 6 mg (2 tablets) | 18 mg per person | $6.00 |
| Coartem® | $3.00 per 24 tablets | A course of 24 tablets per person | $3.00 |

Additionally, each pregnancy test had a unit cost of $0.5 per person. As pregnancy tests were required for female refugees aged between 12 and 45 years old, we estimated that 28.8% of refugees from Africa fell within this demographic and required pregnancy tests in the program. Thus, the estimated operational cost per person was determined to be $10.24 ($10.1+$0.5×28.8%).

To account for overhead costs, we included 7% of the combined staff and operational costs. The estimated average staff and operational costs per person was $27.4. Accordingly, the estimated overhead costs were calculated to be $1.92 per person ($27.4 × 7%). Taking these factors into consideration, the estimated total cost of the overseas presumptive treatment program per U.S.-bound refugees in Africa was $29.32 per person. We assumed a range of ±25% of the baseline values for the lower and upper bound estimates (range: $21.99 - $36.65 per person).

Domestic Presumptive Treatment Costs

The costs for “Domestic Presumptive Treatment” were estimated using parameter estimates already defined in “Domestic Screening and Treatment”. Specifically, we used the same assumption that 10% of the cost of the comprehensive exam could be attributable to presumptive treatment for intestinal parasites and malaria, and the remaining 90% could be used for other health conditions in refugees. We also used the same assumptions regarding drug costs as reported above for individuals treated after diagnosis. The drug regimen for trichuriasis following diagnosis is different than that used for hookworm or ascariasis or that used for “Overseas Presumptive Treatment”. We used the same albendazole regimen as that used for “Overseas Presumptive Treatment”. The costs for a portion of the comprehensive exam, translation, and drug costs are summarized in Table S19. The total cost estimate is $491, range: $392 to $1,086.

Table S19. Estimated costs for Domestic Presumptive Treatment

|  |  | CPT code | No. of tests or visits | Baseline (USD) | Lower bound (USD) | Upper bound (USD) |
| --- | --- | --- | --- | --- | --- | --- |
| Comprehensive exam (new patient, level 4 complexity) | MarketScan® | 99204 | 0.1^a^ | 189 | 156 | 219 |
|  | CMS |  |  | 137 | 137 | 137 |
|  | Weighted |  |  | 150 | 142 | 158 |
|  | (25% × MarketScan® + 75% × CMS) |  |  |  |  |  |
| Translator (0.25 hours × $37.35/hour) | NA | NA | 1 | 9 | 9 | 9 |
| Albendazole (2 × 200mg) for *Ascaris, Trichuris,* or hookworm | NA | NA | NA | 136 | 38 | 582 |
| Praziquantel (3 × 600mg) for *Schistosoma* | NA | NA | NA | 187 | 187 | 299 |
| Ivermectin (2 days × 3 × 3mg) for *S. stercoralis* | NA | NA | NA | 21 | 21 | 33 |
| Coartem® (6 doses × 4 tablets, Tablet=20mg artemether/ 120 mg lumefantrine) for malaria | NA | NA | NA | 123 | 123 | 147 |
| Total cost | NA | NA | NA | 491 | 392 | 1,086 |

### Quality-adjusted life year (QALY) decrement

For uninfected individuals, we used nationally representative QALY values for the non-institutionalized U.S. population from the literature (3). The authors reported estimates for men and women in 10-year age increments using two survey instruments, the health state short form-6 dimensions (SF-6D) and Quality of Well-Being (QWB) Scale (3). The data were collected as part of the 2011 Medical Expenditures Panel Survey and the 2011 National Health Interview Survey. Their results are summarized in Table S20 below based on the mean values for each age group after averaging across men and women and the two survey instruments (SF-6D and QWB Scale).

Table S20: Average QALY weight by age group for a nationally representative sample of the non-institutionalized U.S. population

| Age group | Average QALY weight estimates |
| --- | --- |
| 20-29 | 0.81625 |
| 30-39 | 0.8035 |
| 40-49 | 0.7865 |
| 50-59 | 0.768 |
| 60-69 | 0.75625 |
| 70-79 | 0.73675 |
| 80-89 | 0.689 |

Very limited data are available to estimate the number of QALYs lost to either chronic or acute infections with these parasites in the United States. Expanding our search to the international estimates of QALY weights for these diseases yielded little information. One estimate of QALY weights associated with malaria illness was available from the United States. However, the authors did not interview malaria patients, and only included 11 malaria experts, who estimated that individuals with severe malaria would have a QALY weight of 0.091 for 6.36 days of hospitalization while individuals with simple malaria would have a QALY weight of 0.539 for 3.23 days of hospitalization (64). The authors simply assumed a zero quality of life QALY weight during hospitalization and no disutility (QALY weight = 1.0) before and after the average duration of hospitalization. The authors then reported a one-week QALY score using these assumptions. In comparison, the authors reported a QALY score of 0.9993 for individuals using malaria chemoprophylaxis without adverse events or 0.9802 in a pooled analysis for those experiencing mild adverse effects. Both scores are generally higher than the baseline values for the U.S. population reported above. Given the limited sample size and nature of the survey, we decided instead to consider decrements in QALY values from the baseline values reported in Table S21.

Since these parasitic diseases are more frequently reported in countries that use disability adjusted life years (DALYs) than in countries that use QALYs, we referred to standardized values reported by the WHO (65) as a starting point for measuring QALYs for individuals infected with parasitic diseases. This source provided four relevant estimates of DALY weights: a) Intestinal nematode infections: symptomatic: 0.027, b) Infectious disease: acute episode, mild: 0.006, c) Infectious disease: acute episode, moderate: 0.051, and d) Infectious disease: acute episode, severe: 0.133 (65). Notably, WHO did not report a separate estimate for disutility associated with malaria or schistosomiasis. In addition, WHO only reported a value for acute episodes of nematode infections (hookworm, strongyloidiasis, ascariasis, and trichuriasis).

With this limited information, we attempted to use conservative assumptions to estimate the potential decrements to QALY weights as summarized in Table S21. These decrements would be subtracted from the age-specific QALY weights shown in Table S20. For asymptomatic or subclinical disease, we assumed a value of 0.001 for each parasitic infection as was used in a previous analysis (1) with the exception of individuals with schistosomiasis. This value is approximately 3.7% of the WHO-recommended DALY weights for symptomatic nematode infections and about 16.7% of the value used for mild acute episodes of infectious disease. In comparison to acute episodes, individuals with chronic parasitic infections would not clear the infection within the one-year period (as would be assumed for an individual with mild infectious disease for the WHO recommended value) but would instead continue to be infected over the entire period.

We used a slightly higher value for individuals with schistosomiasis because even chronic light infections have been reported to cause disutility. Authors of one systematic review suggested the use of a QALY weight equal to 0.986 for individuals with light infection (compared to 1.0 for uninfected individuals) (66). This would be equivalent to a decrement of 0.014, which would be 14 times higher than the values we applied for other parasitic infections (66). However, the authors were primarily examining the impact of programs targeted for children and most U.S.-bound refugees are adults. To be conservative, we applied a value of 0.006, which was equivalent to the WHO recommended DALY estimate for individuals with an acute episode of mild infectious disease (65).

For individuals hospitalized with either strongyloidiasis or malaria, we used a different set of assumptions based on the average duration of hospitalization for each disease. For malaria, the average length of stay for U.S. patients was 5.4 days for patients with private insurance and 10 days for patients with Medicaid (61). Thus, we assumed an average of eight days of severe illness with QALY decrement of 0.133 (based on the WHO-recommended value for severe disease) (65). We also assumed that recovery after hospital discharge may include another five days of moderate illness and again applied the WHO-recommended value for moderate disease 0.051 (65). After multiplying the number of days by the DALY weights, we divided by 365 days to obtain a QALY decrement estimate of 0.0036. We applied the same approach for patients hospitalized by strongyloidiasis for which average duration of hospitalization was 13 days (34). We again assumed five days of moderate disease for individuals hospitalized with strongyloidiasis. The resulting QALY decrement is 0.0054. These estimates should be conservative since we only applied the duration of severe and moderate illness to a few days per illness.

Table S21: Summary of estimated QALY decrements for each infection

| Description | Baseline | Lower bound | Upper bound | Distri-bution ^a^ | Ref |
| --- | --- | --- | --- | --- | --- |
| QALY decrement for hookworm, *Ascaris*, *Trichuris* infections | 0.001 | 0 | 0.01 | B | (1) |
| QALY decrement for asymptomatic *Schistosoma* infections | 0.006 | 0.001 | 0.014 | B | (1, 65, 66) |
| QALY decrement for asymptomatic *S. stercoralis* infections | 0.001 | 0 | 0.01 | B | (1) |
| QALY decrement for asymptomatic malaria infection or outpatient malaria, note duration of infection is <1 year | 0.001 | 0 | 0.01 | B | Assumption, (1) |
| QALY decrement for hospitalized malaria | 0.0036 | 0.0018 | 0.0072 | B | (61, 65) |
| QALY decrement for hospitalized strongyloidiasis patient | 0.0054 | 0.0027 | 0.0109 | B | (34, 65) |

^a^ Distribution types: B- generalized beta

**Summary of input parameter estimates**

A summary table of all input parameter estimates is included in Table S22 below.

Table S22. Summary of key epidemiologic, economic, and demographic parameter estimation

| *Study population* | - Average annual cohort of 15,656 African refugees based on Department of Homeland Security data for 2013-2022, median value and average age is 25 years. [Table S8] (39) - We assumed that 90% of these refugees would be covered by presumptive treatment programs. |
| --- | --- |
| *Epidemiologic parameter estimation* | - Test sensitivities for infections vary from 78% for hookworm to 99.5% for malaria (polymerase chain reaction test, PCR or three thick/thin blood smears) based on two stool ova and parasite tests for hookworm, *Ascaris, Trichuris*, one-each *Strongyloides* and *Schistosoma* serologic tests and either three thick/thin blood smear or a single PCR test for malaria. [Table S1] (1, 9-11, 13) - The estimated specificities were 100%, except for *Strongyloides* and *Schistosoma* serologic tests (97% and 84% respectively). [Table S1] (1, 10, 12, 13) - Treatment efficacy in the United States [Table S1]   - Albendazole effectiveness was estimated based on a meta-analysis with reported efficacy varying between 32% against *Trichuris* to 97% against *Ascaris* infections. [Table S2] (4)   - Praziquantel effectiveness was estimated based on a meta-analyses that reported efficacy of about 78% against *Schistosoma mansoni* and 62% against *Schistosoma haemotobium* infections (midpoint value used). (5, 6)   - Ivermectin efficacy was estimated to be 84% against *Strongyloides stercoralis* for a 2-day treatment regimen. (7)   - Artemether-lumefantrine efficacy was estimated to be 97% against malaria. (8) - Overseas presumptive treatment efficacy was assumed to be 95% of these estimates to account for the possibility of re-infection prior to departure and to account for any variations in drug efficacy. - Prevalence   - The prevalence of hookworm, *Trichuris*, and *Ascaris* infections (1.8% to 10.6%) was estimated from a multiyear study of newly-arrived refugees conducted in the State of Minnesota after adjustment for presumptive treatment and test sensitivity (14) using the same approach from a previous analysis. [Table S3] (1)   - The prevalence of schistosomiasis (9.1%) and strongyloidiasis (10.9%) were estimated from a systematic review of rates in migrants. [Table S1] (15)   - The prevalence of malaria (7.2%) was estimated from several published studies focusing on immigrant populations coming from sub-Saharan Africa and living in a country not endemic for malaria for less than one year. (16-22, 24) [Tables S1 and S4] The prevalence estimate was further adjusted for potential differences between U.S. immigrant and refugee populations using data for reported differences in tuberculosis prevalence rates. [Table S5] (46) - The duration of infection in the absence of treatment was estimated to be: Ascariasis one year, Trichuriasis two years, Schistosomiasis five years, hookworm six years, malaria one year, and Strongyloidiasis indefinite. (25-29) [Table S1] - The annual probability of outpatient treatment (0.001) for hookworm, trichuriasis, schistosomiasis, ascariasis, and strongyloidiasis and inpatient treatment (2.9 × 10^-5^ for strongyloidiasis-only) was taken from a previous study (1), which used data from two earlier studies that estimated the incidence of inpatient and outpatient strongyloidiasis among immigrant populations in New York state and Barcelona, Spain. [Table S1] (32, 33) Given a lack of data for each infection, we assumed the annual probability of seeking treatment for each intestinal parasite infection would be the same. This is because strongyloidiasis was frequently diagnosed after an incidental finding of unexplained eosinophilia in patients. (33) - The probability of seeking treatment for malaria was calculated from the baseline prevalence of malaria infection, the reported numbers of malaria outpatient and inpatient cases in the United States, and the number of African immigrants to the United States in 2018. The rates were then adjusted for potential differences between immigrants and refugees based on differences in rates observed for infectious tuberculosis between immigrants and refugees [Tables S1 and S5] (18, 22, 31, 35, 37-39, 46) - The risk of death from inpatient strongyloidiasis was estimated to be 16.7%. [Table S1] (34) - The risk of death from inpatient malaria (0.72%) was estimated from U.S. surveillance data. [Tables S1 and S6] (36, 40) - We assumed that side effects of presumptive treatment would be minor and not of economic significance. |
| *Cost analysis* | - Domestic screening was assumed to include two stool ova and parasite tests, one each serologic test for *Strongyloides* and *Schistosoma* infection, and either a thick/thin blood smear test or PCR test. We also assumed this testing would comprise 10% of a comprehensive examination at a total cost of $102. [Table S9] Unit costs were estimated using two sets of reimbursement rates: 1) 2021 Medicare Clinical Laboratory Fee Schedule or the facility prices of the 2021 Medicare Physician Fee Schedule (MPFS) from the Centers for Medicare and Medicaid Services (CMS) (48, 49) or 2) payments for patients with private insurance using the Merative® MarketScan® Commercial Database (Merative®, Ann Arbor, MI) using the 10 million subsample Set B from January 1, 2021, to December 31, 2021, with an online tool called MarketScan® Treatment Pathways. [Table S10] - Persons with positive test results required a follow-up visit and antiparasitic treatment, for which the cost varies by the type of treatment received from $70 for strongyloidiasis to $458 for trichuriasis. [Table S11] - Medicine costs were estimated from Medicaid, an online pharmacy, and Merative Micromedex® RED BOOK® data (51-54, 56, 63), using the recommended regimen for each infection (41-45, 55). [Table S11] - Outpatient treatment costs for patients treated outside the context of diagnoses during post-arrival medical examinations were based on previously published studies. Cost estimates varied from $340 for ascariasis to $911 for malaria. [Table S13] (34, 59, 60) - Strongyloidiasis ($181,000) and malaria ($20,400) hospitalization costs were also estimated from previously published studies. [Table S14] (34, 58, 61) - Opportunity costs for screening (total: 1.2 hours including time to provide stool and blood samples) and for treatment of individuals diagnosed during screening (1 hour) were estimated using the average U.S. GDP per capita ($8.01 per hour) (57) and the average hourly wage ($28 per hour) (50). [Table S12] - Overseas presumptive treatment costs were estimated in collaboration with IOM based on 2023 data and included medicine, delivery, administrative, and overhead cost data. [Tables S17-S18] - Domestic presumptive treatment costs were estimated to be $491 for the same four-drug regimen as used for overseas presumptive treatment. [Table S19]. |

**Distributions**

We employed distributions for each parameter to conduct probabilistic sensitivity analyses (PSA). The PSAs were conducted using @RISK software version 7, an add-in tool for Microsoft Excel.

Generalized beta distribution

The density function of the generalized beta distribution is shown below.

$$f\left( x \right)=\frac{({x-lower bound)}^{a_{1}-1}{(upper bound-x)}^{a_{2}-1}}{B(a_{1},a_{2}){(upper bound-lower bound)}^{{(a}_{1}+a_{2}-1)}}$$

where x is a continuous variable between lower and upper bound estimates.

The mean and variance of the generalized beta distribution are derived from the following equations.

$$Mean=lower bound+\frac{a_{1}}{a_{1}+a_{2}}\times(upper bound-lower bound)$$

$$Variance=\frac{a_{1}a_{2}}{{{({a_{1}+a}_{2})}^{2}(a}_{1}+a_{2}+1)}\times{(upper bound-lower bound)}^{2}$$

The software, @RISK, requires values for a_1_, a_2_, and upper and lower bound estimate to simulate a generalized beta distribution. To estimate a_1_ and a_2_, we assumed that the mean of the generalized beta distribution corresponds to a baseline value for each parameter. Additionally, we assumed that the standard deviation (SD), which is the squared root of the variance, estimated from the range between the upper and lower bounds divided by 4, using the range rule of thumb. Based on this assumption, we derived the estimates for a_1_ and a_2_ as follows.

$a_{1}=16A^{2}\left( 1-A \right)-A$

$a_{2}=16{(1-A)}^{2}A-1+A$

where $A=\frac{a_{1}}{a_{1}+a_{2}}=\frac{(Mean-lower bound)}{(upper bound-lower bound)}$

Gamma distribution

The density function of the gamma distribution is shown below.

$$f\left( x \right)=x^{(a-1)}{(1-x)}^{(b-1)}\frac{\Gamma(a+b)}{\Gamma(a)\Gamma(b)}$$

where 0<x<1, a>0, and b>0.

The @RISK software requires values for a and b to simulate a gamma distribution. To estimate a and b, we assumed that the mean of the gamma distribution represents a baseline value for each parameter. We also assumed that SD was estimated using the range rule of thumb. Under this assumption, we calculated the estimates for a and b as follows.

$$a=\frac{{Mean}^{2}}{{SD}^{2}}$$

$$b=\frac{{SD}^{2}}{Mean}$$

Uniform distribution

The density function of the uniform distribution is shown below.

$$f\left( x \right)=\frac{1}{(upper bound-lower bound)}$$

where lower bound≤ x ≤upper bound.

### Two-way sensitivity analysis (QALY and incidence multipliers)

To account for the significant uncertainty in prevalence and QALY decrement estimates, we conducted two-way sensitivity analyses in which these parameter estimates were varied by a constant multiplier of between zero and 1.8 for QALY decrements and between 0.2 and 1.8 for prevalence estimates. We applied the same multiplier to all six parasitic diseases simultaneously in contrast to the one-way sensitivity analyses that varied each disease-specific parameter one at a time.

Table S23a shows the expected ICER estimates as functions of multipliers used to adjust parasite prevalence and QALY decrements for infected refugees (holding all other parameters at baseline values) for the comparison between “Overseas Presumptive Treatment” and “No Program”. Relative to the values presented Table 1 and Table S1, the prevalence and QALY decrements for each infection are reduced by the same percentage such that the ICER estimate is greatest (i.e., least cost-effective) when the QALY multiplier is zero and the prevalence multiplier is 0.2. “Overseas Presumptive Treatment” would be cost-saving for prevalence multipliers of 1.0 or greater. With a QALY multiplier of 0, the cost per QALY gained remained below $100,000 for prevalence multipliers greater than or equal to 0.4. By setting the QALY multiplier to zero, only the cost per discounted life year gained is presented (i.e. assuming no disability for infection, outpatient, or inpatient illness). For all observations in Table 3a with a QALY multiplier ≥ 0.2 and a prevalence multiplier ≥ 0.4, the net cost per QALY gained is less than $50,000.

In comparison, “Domestic Screening and Treatment” would not be cost-saving even with prevalence multipliers of 1.8 times the baseline values (Table S23b). With a QALY multiplier of zero, the cost per QALY gained remained at least $270,000 even for a prevalence multiplier of 1.8. When both the prevalence multiplier and QALY multiplier are ≥ 0.8, the cost per QALY gained would be < $100,000; when QALY and prevalence multipliers are both ≥ 1.0, the cost per QALY gained would be less than $50,000.

Finally, “Domestic Presumptive Treatment” required the highest prevalence and QALY multipliers for the alternative to be cost-effective relative to “No Program” (Table S23c). With a QALY multiplier of zero, the cost per QALY gained would be at least $460,000 even with a prevalence multiplier of 1.8. To achieve cost per QALY gained estimates < $100,000, prevalence and QALY multipliers must both be ≥ 1.0; for cost per QALY gained estimates < $50,000, prevalence and QALY multipliers must both be ≥ 1.4.

Table S23a. Two-way sensitivity analyses of infection prevalence and QALY decrement on the net cost per QALY gained, “Overseas Presumptive Treatment” compared to “No Program” (2021 USD)

|  | Prevalence multiplier | | | | | | | | |
| --- | --- | --- | --- | --- | --- | --- | --- | --- | --- |
| QALY multiplier | 0.2 | 0.4 | 0.6 | 0.8 | 1 | 1.2 | 1.4 | 1.6 | 1.8 |
| 0 | $364,805 | $126,673 | $47,296 | $7,607 | Cost-saving | | | | |
| 0.2 | $124,407 | $43,199 | $16,129 | $2,594 |  |  |  |  |  |
| 0.4 | $74,991 | $26,039 | $9,722 | $1,564 |  |  |  |  |  |
| 0.6 | $53,671 | $18,637 | $6,958 | $1,119 |  |  |  |  |  |
| 0.8 | $41,791 | $14,511 | $5,418 | $871 |  |  |  |  |  |
| 1 | $34,216 | $11,881 | $4,436 | $713 |  |  |  |  |  |
| 1.2 | $28,966 | $10,058 | $3,755 | $604 |  |  |  |  |  |
| 1.4 | $25,113 | $8,720 | $3,256 | $524 |  |  |  |  |  |
| 1.6 | $22,165 | $7,696 | $2,874 | $462 |  |  |  |  |  |
| 1.8 | $19,836 | $6,888 | $2,572 | $414 |  |  |  |  |  |

Table S23b. Two-way sensitivity analyses of infection prevalence and QALY decrement on the net cost per QALY gained, “Domestic Screening and Treatment” compared to “No Program” (2021 USD)

|  | Incidence multiplier | | | | | | | | |
| --- | --- | --- | --- | --- | --- | --- | --- | --- | --- |
| QALY multiplier | 0.2 | 0.4 | 0.6 | 0.8 | 1 | 1.2 | 1.4 | 1.6 | 1.8 |
| 0 | $1,845,242 | $959,483 | $664,229 | $516,603 | $428,027 | $368,976 | $326,797 | $295,163 | $270,559 |
| 0.2 | $667,025 | $346,837 | $240,108 | $186,744 | $154,725 | $133,379 | $118,132 | $106,697 | $97,803 |
| 0.4 | $407,091 | $211,678 | $146,540 | $113,971 | $94,430 | $81,402 | $72,097 | $65,118 | $59,690 |
| 0.6 | $292,936 | $152,320 | $105,448 | $82,012 | $67,950 | $58,576 | $51,880 | $46,858 | $42,952 |
| 0.8 | $228,782 | $118,961 | $82,354 | $64,051 | $53,069 | $45,747 | $40,518 | $36,596 | $33,545 |
| 1 | $187,679 | $97,589 | $67,559 | $52,544 | $43,535 | $37,529 | $33,239 | $30,021 | $27,518 |
| 1.2 | $159,097 | $82,726 | $57,270 | $44,541 | $36,904 | $31,813 | $28,176 | $25,449 | $23,328 |
| 1.4 | $138,069 | $71,793 | $49,701 | $38,654 | $32,027 | $27,608 | $24,452 | $22,085 | $20,244 |
| 1.6 | $121,951 | $63,412 | $43,899 | $34,142 | $28,288 | $24,385 | $21,598 | $19,507 | $17,881 |
| 1.8 | $109,203 | $56,783 | $39,310 | $30,573 | $25,331 | $21,836 | $19,340 | $17,468 | $16,012 |

Table S23c. Two-way sensitivity analyses of infection prevalence and QALY decrement on the net cost per QALY gained, “Domestic Presumptive Treatment” compared to “No Program” (2021 USD)

|  | Incidence multiplier | | | | | | | | |
| --- | --- | --- | --- | --- | --- | --- | --- | --- | --- |
| QALY multiplier | 0.2 | 0.4 | 0.6 | 0.8 | 1 | 1.2 | 1.4 | 1.6 | 1.8 |
| 0 | $5,012,711 | $2,451,099 | $1,597,228 | $1,170,293 | $914,131 | $743,357 | $621,376 | $529,890 | $458,734 |
| 0.2 | $1,709,951 | $836,126 | $544,851 | $399,214 | $311,831 | $253,576 | $211,966 | $180,758 | $156,485 |
| 0.4 | $1,030,788 | $504,031 | $328,446 | $240,653 | $187,977 | $152,860 | $127,777 | $108,964 | $94,332 |
| 0.6 | $737,762 | $360,748 | $235,077 | $172,242 | $134,540 | $109,406 | $91,453 | $77,988 | $67,516 |
| 0.8 | $574,459 | $280,897 | $183,043 | $134,116 | $104,760 | $85,189 | $71,210 | $60,726 | $52,571 |
| 1 | $470,347 | $229,989 | $149,869 | $109,810 | $85,774 | $69,750 | $58,304 | $49,720 | $43,043 |
| 1.2 | $398,183 | $194,702 | $126,875 | $92,962 | $72,614 | $59,048 | $49,359 | $42,092 | $36,439 |
| 1.4 | $345,217 | $168,803 | $109,998 | $80,596 | $62,955 | $51,194 | $42,793 | $36,493 | $31,592 |
| 1.6 | $304,688 | $148,985 | $97,084 | $71,134 | $55,564 | $45,184 | $37,769 | $32,208 | $27,883 |
| 1.8 | $272,675 | $133,332 | $86,884 | $63,660 | $49,726 | $40,436 | $33,801 | $28,824 | $24,954 |

Note: Net costs did not include illness opportunity costs but did include intervention opportunity costs.

### Additional Results Tables

Table S24. Expanded results with outcomes reported by parasitic disease (N=15,656)

|  | “No Program” | “Domestic Screening and Treatment” | “Overseas Presumptive Treatment” | “Domestic Presumptive Treatment” |
| --- | --- | --- | --- | --- |
| Fraction domestic screen/treat | 0% | 90% | 9% | 9% |
| Fraction overseas presumptive treatment | 0% | 0% | 90% | 0% |
| Fraction domestic presumptive treatment | 0% | 0% | 0% | 81% |
| Fraction no program | 100% | 10% | 1% | 10% |
| Overseas or domestic presumptive treatment or domestic screening (not including domestic treatment for those diagnosed with infection) | | | | |
| Overseas or domestic presumptive treatment costs | $0 | $0 | $412,841 | $6,226,428 |
| Domestic screening costs | $0 | $1,437,193 | $143,719 | $143,719 |
| Opportunity costs for domestic screening | $0 | $140,901 | $14,090 | $14,090 |
| *Subtotal presumptive treatment and domestic screening costs* | *$0* | *$1,578,095* | *$570,650* | *$6,384,238* |
| Costs to treat individuals diagnosed with each infection during domestic screening | | | | |
| Hookworm | $0 | $36,598 | $3,660 | $3,660 |
| Ascariasis | $0 | $223,809 | $22,381 | $22,381 |
| Trichuriasis | $0 | $477,026 | $47,703 | $47,703 |
| Strongyloidiasis | $0 | $125,271 | $12,527 | $12,527 |
| Schistosomiasis | $0 | $731,759 | $73,176 | $73,176 |
| Malaria | $0 | $173,620 | $17,362 | $17,362 |
| *Subtotal treatment costs for individuals identified during domestic screening* | *$0* | *$1,768,083* | *$176,808* | *$176,808* |
| Opportunity costs for treating individuals diagnosed during domestic screening | | | | |
| Hookworm | $0 | $1,583 | $158 | $158 |
| Ascariasis | $0 | $9,678 | $968 | $968 |
| Trichuriasis | $0 | $8,332 | $833 | $833 |
| Strongyloidiasis | $0 | $14,317 | $1,432 | $1,432 |
| Schistosomiasis | $0 | $24,805 | $2,481 | $2,481 |
| Malaria | $0 | $8,075 | $808 | $808 |
| *Subtotal opportunity costs for individuals treated after diagnosis during domestic screening* | *$0* | *$66,791* | *$6,679* | *$6,679* |
| **Total intervention costs** | **$0** | **$3,412,968** | **$754,138** | **$6,567,725** |
| Cases diagnosed after completion of interventions | | | |  |
| Total outpatient cases (not discounted) | | | | |
| Hookworm | 1.7 | 0.8 | 0.5 | 0.5 |
| Ascariasis | 1.7 | 0.5 | 0.2 | 0.2 |
| Trichuriasis | 2.4 | 1.7 | 1.7 | 1.7 |
| Strongyloidiasis | 84.8 | 25.8 | 18.0 | 21.2 |
| Schistosomiasis | 7.1 | 3.4 | 2.5 | 2.7 |
| Malaria | 16.6 | 2.2 | 1.4 | 2.1 |
| *Total outpatient cases* | *114* | *34* | *24* | *28* |
| Total hospitalized cases (not discounted) | | | | |
| Hookworm | 0.0 | 0.0 | 0.0 | 0.0 |
| Ascariasis | 0.0 | 0.0 | 0.0 | 0.0 |
| Trichuriasis | 0.0 | 0.0 | 0.0 | 0.0 |
| Strongyloidiasis | 2.5 | 0.7 | 0.5 | 0.6 |
| Schistosomiasis | 0.0 | 0.0 | 0.0 | 0.0 |
| Malaria | 32.0 | 4.2 | 2.7 | 4.1 |
| *Total hospitalized cases* | *34.48* | *4.96* | *3.20* | *4.70* |
| Deaths (not discounted) | | | | |
| Hookworm | N/A | N/A | N/A | N/A |
| Ascariasis | N/A | N/A | N/A | N/A |
| Trichuriasis | N/A | N/A | N/A | N/A |
| Strongyloidiasis | 0.411 | 0.125 | 0.087 | 0.103 |
| Schistosomiasis | N/A | N/A | N/A | N/A |
| Malaria | 0.231 | 0.030 | 0.019 | 0.029 |
| *Total deaths* | *0.641* | *0.155* | *0.107* | *0.132* |
| Total QALYs lost (discounted) | | | | |
| Hookworm | 1.54 | 0.69 | 0.42 | 0.48 |
| Ascariasis | 1.64 | 0.49 | 0.17 | 0.24 |
| Trichuriasis | 2.34 | 1.69 | 1.63 | 1.67 |
| Strongyloidiasis | 45.91 | 13.98 | 9.74 | 11.48 |
| Schistosomiasis | 39.53 | 19.11 | 13.83 | 15.07 |
| Malaria | 5.72 | 0.75 | 0.48 | 0.73 |
| *Total QALYs lost* | *96.68* | *36.70* | *26.28* | *29.67* |
| Lifetime discounted illness treatment costs (after intervention) | | | |  |
| Hookworm | $600 | $269 | $164 | $185 |
| Ascariasis | $554 | $164 | $58 | $82 |
| Trichuriasis | $1,853 | $1,339 | $1,293 | $1,320 |
| Strongyloidiasis | $248,667 | $75,714 | $52,779 | $62,179 |
| Schistosomiasis | $5,569 | $2,692 | $1,948 | $2,124 |
| Malaria | $657,728 | $86,402 | $55,109 | $83,819 |
| *Total discounted illness treatment costs* | *$914,971* | *$166,581* | *$111,351* | *$149,708* |
| Lifetime discounted illness opportunity costs (after intervention) | | | | |
| Hookworm | $295 | $132 | $81 | $91 |
| Ascariasis | $313 | $93 | $33 | $46 |
| Trichuris | $448 | $323 | $312 | $319 |
| Strongyloides | $11,264 | $3,430 | $2,391 | $2,817 |
| Schistosomiasis | $1,261 | $609 | $441 | $481 |
| Malaria | $51,522 | $6,768 | $4,317 | $6,566 |
| *Total discounted illness opportunity costs* | *$65,103* | *$11,356* | *$7,574* | *$10,319* |

N/A; Not available

**Additional Results Figures**

Figure S4a. One-way sensitivity analysis of net cost per QALY gained for “Overseas Presumptive Treatment” vs. “No Program”, all parameters causing the greatest difference in net cost per QALY gained estimates between lower and upper bounds, baseline value: -$1,520 2021 USD ^a^

Net cost per QALY gained, 2021 USD

|  | Lower bound | Upper bound |
| --- | --- | --- |
| QALY decrement: strongyloidiasis- subclinical/oupatient | 88891.1 | 7784.83 |
| Strongyloidiasis prevalence | 76092.5 | 25475.2 |
| Schistosomiasis Prevalence | 60027.8 | 27779.8 |
| QALY decrement: schistosomiasis- subclinical/oupatient | 60781.6 | 29941.1 |
| Cost: trichuriasis tx after positive test | 38185.5 | 66806.8 |
| Inf Duration: schistosomiasis | 50044.1 | 33718.7 |
| Annual Prob inpatient strongyloidiasis tx | 46874.2 | 31707.4 |
| Malaria prevalence | 47735 | 33705.8 |
| Annual Prob inpatient tx malaria | 49936.1 | 35988.3 |
| Cost: hookworm or ascariasis tx after positive test | 41164 | 54049.7 |
| Cost: inpatient tx malaria | 48872.4 | 38197.3 |
| QALY decrement: hookworm, ascariasis, trichuriasis- subclinical/oupatient | 44881.2 | 34278.5 |
| Ivermectin efficacy against strongyloidiasis | 47584.6 | 39524.8 |
| Trichuriasis prevalence | 39708.1 | 47320 |
| Screen cost for all parasites | 39305.5 | 46119 |
| QALY decrement: malaria subclinical/oupatient | 44233.9 | 38112 |
| Cost: schistosomiasis tx after positive test | 43431.2 | 49428.7 |
| Opp cost: screening (all parasites) | 43534.6 | 49173.3 |
| Case fatality rate for inpatient strongyloidiasis | 44323.5 | 39461.5 |
| Specificity: Schistosomiasis ELISA test | 46140.1 | 41450.2 |
| Praziquantel efficacy against Schistosoma mansoni | 46485.1 | 41799.2 |
| Cost: inpatient tx strongyloidiasis | 45822.8 | 41778.3 |
| Case fatality rate for inpatient malaria | 44505.7 | 40514.2 |
| Ascariasis prevalence | 41995.9 | 45044 |
| Praziquantel efficacy against Schistosoma haemotobium | 45304.3 | 42295.1 |
| Opp cost: tx after screening | 43534.6 | 46318.9 |
| Sesnsitivity: schistosomiasis ELISA test | 45167.9 | 42533.2 |
| Sesnsitivity: strongyloidiasis serologic test | 44618.6 | 42248.1 |
| Sesnsitivity: two stool O&P for ascariasis | 42626.4 | 44059.6 |
| Sesnsitivity: two stool O&P for trichuiasis | 42343.3 | 43772.4 |
| Inf Duration: ascariasis | 43534.6 | 42730.9 |
| Inf Duration: hookworm | 44052 | 43442.1 |
| Cost: strongyloidiasis tx after positive test | 43445.1 | 43952.4 |
| Specificity: strongyloidiasis serologic test | 43697.9 | 43208 |
| Annual Prob outpatient tx hookworm/trichuriasis/ascariasis | 43451.6 | 43923.5 |
| Cost: malaria tx after positive test | 43500.9 | 43972.2 |
| Opp cost: inpatient malaria | 43797.4 | 43417.8 |
| Albendazole efficacy against trichuiasis | 43668.5 | 43379.1 |
| Coartem efficacy against malaria | 43677.8 | 43391.6 |
| Inf Duration: trichuiasis | 43772.7 | 43534.6 |
| Annual Prob outpatient tx malaria | 43613.5 | 43422.8 |
| Cost: outpatient illness malaria | 43612.5 | 43456.9 |
| Cost: outpatient illness strongyloidiasis | 43597.8 | 43465.4 |
| Annual Prob outpatient tx strongyloidiasis/schistosomiasis | 43556.5 | 43436.3 |
| Albendazole efficacy against hookworm | 43590.3 | 43489.3 |
| Sesnsitivity: thick/thin blood smear for Plasmodium spp. | 43589.2 | 43491 |
| Opp cost: inpatient strongyloidiasis | 43559.4 | 43522.2 |
| QALY decrement: malaria hospitalized | 43570.5 | 43534.6 |
| Cost: outpatient illness schistosomiasis | 43552.9 | 43518.2 |
| Albendazole efficacy against ascariasis | 43553 | 43522.3 |
| Opp cost: outpatient cases | 43534.6 | 43508 |
| Cost: outpatient illness trichuriasis | 43538.6 | 43519.5 |
| Cost: outpatient illness ascariasis | 43538 | 43524.6 |
| Hookworm prevalence | 43528.2 | 43540.8 |

Figure S4b. One-way sensitivity analysis of net cost per QALY gained for “Domestic Screening and Treatment” vs. “No Program”, all parameters causing the greatest difference in net cost per QALY gained estimates between lower and upper bounds, baseline value: $43,535, 2021 USD

Net cost per QALY gained, 2021 USD

|  | Lower bound | Upper bound |
| --- | --- | --- |
| Malaria prevalence | 1897.65 | -9604.89 |
| Annual Prob inpatient tx malaria | 2875.42 | -6739.63 |
| Cost: inpatient tx malaria | 3276.16 | -6315.83 |
| Annual Prob inpatient strongyloidiasis tx | 455.206 | -8546.21 |
| % refugees who would receive presumptive tx | 2815.87 | -5715.54 |
| Strongyloidiasis prevalence | 1051.06 | -3002.32 |
| Cost: inpatient tx strongyloidiasis | 687.677 | -3214.63 |
| Cost: overseas presumptive tx in Africa | -2983.11 | -48.9708 |
| QALY decrement: strongyloidiasis- subclinical/oupatient | -3007.25 | -278.862 |
| Cost: trichuriasis tx after positive test | -1975.72 | 462.458 |
| Schistosomiasis Prevalence | -2693.59 | -474.099 |
| Annual Prob outpatient tx hookworm/trichuriasis/ascariasis | -1205.19 | -2893.38 |
| QALY decrement: schistosomiasis- subclinical/oupatient | -2184.65 | -1022.39 |
| Cost: hookworm or ascariasis tx after positive test | -1721.99 | -624.286 |
| Adjustment factor for overseas versus domestic tx 0-1 | -983.561 | -2007.24 |
| % refugees receiving domestic comprehensive medical exam | -1950.57 | -1097.59 |
| Trichuriasis prevalence | -1869.2 | -1174.37 |
| Ivermectin efficacy against strongyloidiasis | -1192.01 | -1846.69 |
| Screen cost for all parasites | -1880.3 | -1299.88 |
| Inf Duration: schistosomiasis | -1737.5 | -1200.82 |
| Cost: schistosomiasis tx after positive test | -1528.85 | -1017.94 |
| Opp cost: screening (all parasites) | -1520.04 | -1039.69 |
| Specificity: Schistosomiasis ELISA test | -1298.08 | -1697.61 |
| QALY decrement: hookworm, ascariasis, trichuriasis- subclinical/oupatient | -1568.42 | -1189.77 |
| Ascariasis prevalence | -1698.02 | -1345.73 |
| Opp cost: inpatient malaria | -1283.86 | -1625.01 |
| Annual Prob outpatient tx strongyloidiasis/schistosomiasis | -1463.48 | -1775.23 |
| Opp cost: tx after screening | -1520.04 | -1282.86 |
| Annual Prob outpatient tx malaria | -1439.07 | -1634.79 |
| QALY decrement: malaria subclinical/oupatient | -1541.95 | -1347.74 |
| Coartem efficacy against malaria | -1425.98 | -1613.96 |
| Case fatality rate for inpatient strongyloidiasis | -1546.6 | -1382.37 |
| Sesnsitivity: two stool O&P for ascariasis | -1618.84 | -1462.45 |
| Praziquantel efficacy against Schistosoma mansoni | -1618.03 | -1462.85 |
| Cost: outpatient illness malaria | -1450.05 | -1589.82 |
| Sesnsitivity: two stool O&P for trichuiasis | -1627.86 | -1498.48 |
| Cost: outpatient illness strongyloidiasis | -1459.02 | -1586.79 |
| Case fatality rate for inpatient malaria | -1550.44 | -1424.61 |
| Praziquantel efficacy against Schistosoma haemotobium | -1578.7 | -1479.16 |
| Sesnsitivity: schistosomiasis ELISA test | -1579.34 | -1480.79 |
| Hookworm prevalence | -1554.89 | -1485.74 |
| Cost: strongyloidiasis tx after positive test | -1527.67 | -1484.45 |
| Specificity: strongyloidiasis serologic test | -1506.13 | -1547.86 |
| Cost: malaria tx after positive test | -1522.91 | -1482.76 |
| Cost: outpatient illness schistosomiasis | -1500.45 | -1537.63 |
| Opp cost: inpatient strongyloidiasis | -1496.15 | -1531.99 |
| Opp cost: outpatient cases | -1520.04 | -1545.52 |
| Sesnsitivity: two stool O&P for hookworm | -1537.79 | -1512.95 |
| Inf Duration: ascariasis | -1520.04 | -1500.5 |
| Cost: outpatient illness trichuriasis | -1516.33 | -1534.08 |
| Inf Duration: hookworm | -1532.42 | -1517.83 |
| Cost: outpatient illness ascariasis | -1516.39 | -1530.89 |
| Cost: outpatient illness hookworm | -1518 | -1527.56 |
| Sesnsitivity: thick/thin blood smear for Plasmodium spp. | -1517 | -1522.48 |
| Albendazole efficacy against trichuiasis | -1521.54 | -1518.3 |
| Sesnsitivity: strongyloidiasis serologic test | -1518.68 | -1521.74 |
| Inf Duration: trichuiasis | -1522.7 | -1520.04 |
| Albendazole efficacy against hookworm | -1521.37 | -1518.96 |

Figure S4c. One-way sensitivity analysis of net cost per QALY gained for “Domestic Presumptive Treatment” vs. “No Program”, all parameters between lower and upper bounds, baseline value: $85,774, 2021 USD

Net cost per QALY gained, 2021 USD

|  | Lower bound | Upper bound |
| --- | --- | --- |
| QALY decrement: strongyloidiasis- subclinical/oupatient | 169720 | 15733.9 |
| Cost: domestic presumptive tx | 67076.6 | 198335 |
| Strongyloidiasis prevalence | 146694 | 50658.8 |
| Schistosomiasis Prevalence | 132188 | 44393.3 |
| QALY decrement: schistosomiasis- subclinical/oupatient | 123260 | 57698.2 |
| Inf Duration: schistosomiasis | 99641.2 | 65414.5 |
| QALY decrement: hookworm, ascariasis, trichuriasis- subclinical/oupatient | 88503.2 | 67138.8 |
| Annual Prob inpatient strongyloidiasis tx | 90159.4 | 70169.6 |
| Malaria prevalence | 91698.7 | 71758.6 |
| Annual Prob inpatient tx malaria | 92764 | 77473.2 |
| Ivermectin efficacy against strongyloidiasis | 93016.8 | 78561.5 |
| QALY decrement: malaria subclinical/oupatient | 87010.4 | 76046.6 |
| Praziquantel efficacy against Schistosoma mansoni | 92022.3 | 82126.7 |
| Cost: inpatient tx malaria | 90572.5 | 80975.4 |
| Case fatality rate for inpatient strongyloidiasis | 87272.8 | 78004.2 |
| % refugees who would receive presumptive tx | 81523.9 | 89925.6 |
| Case fatality rate for inpatient malaria | 87490 | 80386.1 |
| Praziquantel efficacy against Schistosoma haemotobium | 89514.4 | 83166.8 |
| Cost: inpatient tx strongyloidiasis | 87981.9 | 84078.9 |
| Cost: trichuriasis tx after positive test | 85295 | 87856.6 |
| Annual Prob outpatient tx hookworm/trichuriasis/ascariasis | 85326.5 | 87773.1 |
| Inf Duration: ascariasis | 85773.8 | 84070.3 |
| Ascariasis prevalence | 86504.6 | 85058 |
| Inf Duration: hookworm | 86912 | 85570.6 |
| Hookworm prevalence | 86435.9 | 85122.1 |
| Cost: hookworm or ascariasis tx after positive test | 85561.6 | 86714.8 |
| Screen cost for all parasites | 85395.3 | 86005.1 |
| Sesnsitivity: schistosomiasis ELISA test | 86102.5 | 85556.3 |
| Cost: schistosomiasis tx after positive test | 85764.5 | 86301.3 |
| Albendazole efficacy against trichuiasis | 86016 | 85492.4 |
| Opp cost: screening (all parasites) | 85773.8 | 86278.4 |
| Inf Duration: trichuiasis | 86204.5 | 85773.8 |
| Specificity: Schistosomiasis ELISA test | 86007 | 85587.2 |
| Sesnsitivity: strongyloidiasis serologic test | 85956.4 | 85546.5 |
| Opp cost: inpatient malaria | 86010.1 | 85668.8 |
| Coartem efficacy against malaria | 85935 | 85612.8 |
| Opp cost: tx after screening | 85773.8 | 86023 |
| Albendazole efficacy against hookworm | 85896.2 | 85674.4 |
| Annual Prob outpatient tx malaria | 85835.4 | 85686.6 |
| Trichuriasis prevalence | 85847.6 | 85700.7 |
| Cost: outpatient illness malaria | 85843.8 | 85704 |
| Cost: outpatient illness strongyloidiasis | 85834.8 | 85707 |
| Sesnsitivity: two stool O&P for trichuiasis | 85673.7 | 85793.8 |
| Sesnsitivity: two stool O&P for ascariasis | 85714.4 | 85808.4 |
| Albendazole efficacy against ascariasis | 85812.9 | 85747.7 |
| QALY decrement: malaria hospitalized | 85837.4 | 85773.8 |
| Cost: strongyloidiasis tx after positive test | 85765.8 | 85811.2 |
| Specificity: strongyloidiasis serologic test | 85788.4 | 85744.6 |
| Annual Prob outpatient tx strongyloidiasis/schistosomiasis | 85766.3 | 85808.8 |
| Cost: malaria tx after positive test | 85770.8 | 85812.9 |
| Cost: outpatient illness schistosomiasis | 85793.4 | 85756.2 |
| Opp cost: inpatient strongyloidiasis | 85797.7 | 85761.8 |
| Opp cost: outpatient cases | 85773.8 | 85748.3 |
| Sesnsitivity: two stool O&P for hookworm | 85790.6 | 85767.1 |
| Cost: outpatient illness trichuriasis | 85777.5 | 85759.7 |
| Cost: outpatient illness ascariasis | 85777.4 | 85762.9 |
| Sesnsitivity: thick/thin blood smear for Plasmodium spp. | 85780.2 | 85768.6 |

^a^ The tornado diagram is a series of one-way sensitivity analyses in which parameters are varied (one at a time across their uncertainty ranges while holding all other parameters at their baseline value).

Tx- Treatment, CFR case fatality rate, QALY- quality adjusted life year, Opp.- opportunity, Inf- infection, Prob- probability, stool O&P- stool ova and parasites test, ELISA- enzyme-linked immunosorbent assay

Figure S5. Fraction of Monte Carlo Simulation iterations in which each intervention would be considered cost-effective as a function of willingness to pay per QALY gained (log scale) for “Overseas Presumptive Treatment”, “Domestic Screening and Treatment”, or “Domestic Presumptive Treatment” relative to “No Program”

Net cost per QALY gained threshold

| Fraction of simulations below net cost per QALY threshold | Domestic Screening and Treatment | Overseas Presumptive Treatment | Domestic Presumptive Treatment |
| --- | --- | --- | --- |
| 1% | 5033.65 | -75995.2 | 8323.23 |
| 2% | 6295.1 | -59225.3 | 11168.9 |
| 3% | 7501.59 | -49408.9 | 13610.2 |
| 4% | 8732.5 | -42137.6 | 15836.7 |
| 5% | 10015.6 | -36934.7 | 18176.2 |
| 6% | 11445.5 | -33262.9 | 20589.5 |
| 7% | 12924.7 | -29762.7 | 23083.8 |
| 8% | 14551.2 | -26975.4 | 25534.9 |
| 9% | 16232.3 | -24460.5 | 28198.1 |
| 10% | 17980.8 | -22431.9 | 30646.4 |
| 11% | 19749.8 | -20594.5 | 33388.6 |
| 12% | 21503.2 | -19002.9 | 35991.7 |
| 13% | 23203.3 | -17512.2 | 38710.7 |
| 14% | 24840.5 | -16253.2 | 41406 |
| 15% | 26624.6 | -14961 | 44104.6 |
| 16% | 28456.5 | -13796.5 | 46846.6 |
| 17% | 30135.5 | -12856.5 | 49665 |
| 18% | 31803 | -11859.8 | 52524.7 |
| 19% | 33406.6 | -10968.1 | 55375.2 |
| 20% | 35134.6 | -10217.6 | 58125.7 |
| 21% | 36728.4 | -9501.42 | 61031.5 |
| 22% | 38486.1 | -8719.93 | 63820.5 |
| 23% | 40131 | -8029.6 | 66901.1 |
| 24% | 41714.9 | -7471.54 | 69896.9 |
| 25% | 43474.8 | -6916.65 | 72951.7 |
| 26% | 45109 | -6354.63 | 75950 |
| 27% | 46853.8 | -5804.7 | 79011.9 |
| 28% | 48525.5 | -5295.37 | 82199.3 |
| 29% | 50412.2 | -4879.06 | 85582.4 |
| 30% | 52069.1 | -4446.41 | 88778.5 |
| 31% | 53822.2 | -4053.23 | 91968.2 |
| 32% | 55638.6 | -3709.73 | 95233.3 |
| 33% | 57371.8 | -3373.3 | 98824.3 |
| 34% | 59172.9 | -3013.33 | 102161 |
| 35% | 61086.5 | -2700.71 | 105458 |
| 36% | 63054.8 | -2402.56 | 109132 |
| 37% | 64852.9 | -2114.55 | 112748 |
| 38% | 66794.1 | -1844.7 | 116755 |
| 39% | 68792.9 | -1600.88 | 120440 |
| 40% | 70947.7 | -1375.84 | 124241 |
| 41% | 72950.2 | -1164.16 | 128221 |
| 42% | 75141.2 | -976.951 | 132130 |
| 43% | 77070.9 | -802.152 | 136427 |
| 44% | 79195.5 | -633.141 | 140415 |
| 45% | 81441.9 | -477.36 | 144804 |
| 46% | 83603.6 | -326.988 | 149220 |
| 47% | 85795.4 | -167.864 | 153564 |
| 48% | 87871.9 | -15.4006 | 158466 |
| 49% | 90062 | 118.78 | 163256 |
| 50% | 92478.6 | 265.475 | 168503 |
| 51% | 94981.4 | 402.593 | 173914 |
| 52% | 97797.2 | 555.37 | 178959 |
| 53% | 100601 | 700.562 | 184335 |
| 54% | 103159 | 861.468 | 190132 |
| 55% | 106001 | 1040.04 | 196128 |
| 56% | 108622 | 1212.25 | 202090 |
| 57% | 111357 | 1401.31 | 208054 |
| 58% | 114410 | 1592.83 | 214041 |
| 59% | 117608 | 1785.29 | 220792 |
| 60% | 120595 | 2003.2 | 227435 |
| 61% | 123800 | 2231.41 | 235069 |
| 62% | 127359 | 2503.84 | 242247 |
| 63% | 130974 | 2776.98 | 249170 |
| 64% | 134820 | 3048.56 | 257262 |
| 65% | 138615 | 3314.94 | 264934 |
| 66% | 142687 | 3609.64 | 272672 |
| 67% | 146917 | 3917.71 | 280837 |
| 68% | 151255 | 4252.1 | 290154 |
| 69% | 155680 | 4601.36 | 300031 |
| 70% | 160122 | 4995.84 | 310759 |
| 71% | 164833 | 5387.94 | 320795 |
| 72% | 169410 | 5837.28 | 332466 |
| 73% | 174518 | 6245.72 | 344511 |
| 74% | 180155 | 6708.14 | 356447 |
| 75% | 186104 | 7192.08 | 368429 |
| 76% | 191750 | 7686.9 | 382131 |
| 77% | 198304 | 8252.63 | 395621 |
| 78% | 204453 | 8872.34 | 409657 |
| 79% | 211903 | 9531.74 | 424936 |
| 80% | 219237 | 10225.6 | 440495 |
| 81% | 227062 | 10998.3 | 458169 |
| 82% | 235444 | 11811.5 | 476965 |
| 83% | 244505 | 12705.4 | 497701 |
| 84% | 255169 | 13710.6 | 518147 |
| 85% | 265035 | 14728.4 | 541497 |
| 86% | 276364 | 15924.4 | 569447 |
| 87% | 289174 | 17384.7 | 599292 |
| 88% | 302489 | 19028.5 | 630516 |
| 89% | 317306 | 20705.4 | 666562 |
| 90% | 333657 | 22727.5 | 703112 |
| 91% | 351043 | 25060.6 | 744953 |
| 92% | 371765 | 27612.2 | 795186 |
| 93% | 394511 | 30851.5 | 849109 |
| 94% | 422070 | 34912.7 | 919788 |
| 95% | 455500 | 39602.8 | 991791 |
| 96% | 497578 | 45475.4 | 1086187 |
| 97% | 551525 | 53271.2 | 1214303 |
| 98% | 634952 | 65604.5 | 1409171 |
| 99% | 772266 | 89847.2 | 1767466 |
| 99.25% | 823339 | 100490 | 1912492 |
| 99.5% | 907123 | 115475 | 2102790 |
| 99.75% | 1046921 | 146530 | 2493447 |
| 99.9% | 1252909 | 188998 | 3051510 |
| 99.99% | 1735699 | 314298 | 4697064 |
| 99.99% | 1735699 | 314298 | 4697064 |

42.	WHO. Assessing the efficacy of anthelminthic drugs against schistosomiasis and soil-transmitted helminthiases. 2013.

52.	Red Book Online. Albenza. In: Truven Health Analytics within Micromedex, editor. Greenwood Village, CO:

; 2021.

53.	Red Book Online. Biltricide. In: Truven Health Analytics within Micromedex, editor. Greenwood Village, CO.:

; 2021.

54.	Red Book Online. Stromectol. In: Truven Health Analytics within Micromedex, editor. Greenwood Village, CO:

; 2021.

55.	CDC. Malaria in the United States: Treatment Tables. 2024 [cited January 24 2025]; Available from: <https://www.cdc.gov/malaria/hcp/clinical-guidance/malaria-treatment-tables.html>

56.	Red Book Online. Coartem. In: Truven Health Analytics within Micromedex, editor. Greenwood Village, CO:

; 2021.

57.	The World Bank Group. GDP per capita (current US$)-United States (2021). 2023 [cited; Available from: <https://data.worldbank.org/indicator/NY.GDP.PCAP.CD?locations=US>

58.	Bureau of Labor Statistics. CPI for All Urban Consumers (CPI-U): Medical care in U.S. city average, all urban consumers, not seasonally adjusted. 2023 [cited 2023 Feburary 17]; Available from: <https://data.bls.gov/cgi-bin/surveymost>

59.	Joo H, Lee J, Maskery BA, Park C, Alpern JD, Phares CR, et al. The Effect of Drug Pricing on Outpatient Payments and Treatment for Three Soil-Transmitted Helminth Infections in the United States, 2010-2017. Am J Trop Med Hyg. 2021 Mar 8;104(5):1851-7.

60.	Joo H, Maskery BA, Alpern JD, Chancey RJ, Weinberg M, Stauffer WM. Low Use of Standard-of-Care Antiparasitic Drugs and Increased Estimated Outpatient Payments for Treating Schistosomiasis in the United States, 2013-19. Am J Trop Med Hyg. 2022;107(4):841-4.

61.	Park J, Joo H, Maskery BA, Alpern JD, Weinberg M, Stauffer WM. Costs of malaria treatment in the United States. J Travel Med. 2023;taad013.
